# Realizing the Potential of Social Determinants Data: A Scoping Review of Approaches for Screening, Linkage, Extraction, Analysis and Interventions

**DOI:** 10.1101/2024.02.04.24302242

**Authors:** Chenyu Li, Danielle L. Mowery, Xiaomeng Ma, Rui Yang, Ugurcan Vurgun, Sy Hwang, Hayoung Kim Donnelly, Harsh Bandhey, Zohaib Akhtar, Yalini Senathirajah, Eugene Mathew Sadhu, Emily Getzen, Philip J Freda, Qi Long, Michael J. Becich

## Abstract

**Background:** Social determinants of health (SDoH) like socioeconomics and neighborhoods strongly influence outcomes, yet standardized SDoH data is lacking in electronic health records (EHR), limiting research and care quality.

**Methods:** We searched PubMed using keywords “SDOH” and “EHR”, underwent title/abstract and full-text screening. Included records were analyzed under five domains: 1) SDoH screening and assessment approaches, 2) SDoH data collection and documentation, 3) Use of natural language processing (NLP) for extracting SDoH, 4) SDoH data and health outcomes, and 5) SDoH-driven interventions.

**Results:** We identified 685 articles, of which 324 underwent full review. Key findings include tailored screening instruments implemented across settings, census and claims data linkage providing contextual SDoH profiles, rule-based and neural network systems extracting SDoH from notes using NLP, connections found between SDoH data and healthcare utilization/chronic disease control, and integrated care management programs executed. However, considerable variability persists across data sources, tools, and outcomes.

**Discussion:** Despite progress identifying patient social needs, further development of standards, predictive models, and coordinated interventions is critical to fulfill the potential of SDoH-EHR integration. Additional database searches could strengthen this scoping review. Ultimately widespread capture, analysis, and translation of multidimensional SDoH data into clinical care is essential for promoting health equity.

## Introduction

The concept of social determinants of health (SDoH) acknowledges that health is not simply a product of biological factors or access to medical care, but is profoundly influenced by the social, economic, and physical conditions that shape people’s lives [1]. Research across disciplines including public health, sociology, economics, and medicine provides clear evidence that circumstances in the environments where people exist have a fundamental impact on shaping patterns of health and well-being [2]. The World Health Organization defines SDoH as “*the conditions in which people are born, grow, live, work, and age, along with the wider set of forces and systems shaping the conditions of daily life”* [3]. These determinants are broadly categorized into five interdependent domains that form the structural and social hierarchies in society: **economic stability, neighborhood and built environment, health care access, education access and quality,** and **social and community context** [4,5]

Specifically, adverse SDoH like poverty, unequal access to education, lack of public resources in neighborhoods, high crime rates, racial segregation, and pollution have all been extensively linked to higher rates of morbidity, mortality, and health risk behaviors across populations [1]. On the other hand, protective and promoting SDoH like higher household income, safe green spaces, strong social support, affordable nutrition options, and accessible transportation demonstrate significant associations with positive health indicators, from self-rated health status to reduced prevalence of diabetes to longer life expectancy.

Health outcomes are significantly influenced by more than just clinical encounters; indeed, research suggests that only about 20% of a person’s health outcomes can be attributed to clinical care [6,7]. The majority is determined by a combination of individual behaviors and various external factors that are collectively referred to as SDoH. These “causes of the causes” of health, or upstream social determinants, are estimated to account for up to 55% of population health variation in high-income countries, though some models suggest they may account for as much as 70-80% [7]. Aspects of physical environment, socioeconomic status, race, and gender contribute to systemic inequities that become embodied as adverse outcomes. This makes social determinants fundamental considerations for achieving health equity and overarching population health improvement [1].

Healthy People 2030 in the United States includes objectives to reduce homelessness, increase educational attainment, and improve access to care – aligning with several SDoH domains [8]. The “Improving Social Determinants of Health Act of 2021”[5] further legislates Centers for Disease Control and Prevention [9].

### SDoH-driven Translational Research: Deriving and Translating Health Data to Actionable Knowledge into Clinical Care

Incorporating SDoH into clinical practice is essential for achieving health equity, yet these determinants are seldom consistently recorded or collected within or outside of electronic health records (EHRs). This lack of information is problematic because healthcare professionals can only address SDoH with interventions and programs effectively if they are aware of and can access this information.

Given the impact of SDoH on health outcomes and the limitations of existing EHRs and documentation practices, public health, care providers, and clinical researchers have implemented various approaches to standardize the collection, study, and mobilization of knowledge generated from this information into clinical care (see **Figure 1**). SDoH data, often collected from the community or public health surveys [10], discrete modules within the EHR [11], and patient-reported outcome surveys [12], can be aggregated into a unified SDoH repository, supporting targeted research initiatives. This integration transforms scattered SDoH data into a structured resource for comprehensive study. However, the collection, integration, and utility of this data remain varied and inconsistent across systems. To be useful, this information must be not just integrated, but also standardized using SDoH ontology [13], common representations [14], and value sets [15]. Once integrated and standardized, SDoH information can be studied in association with positive and negative patient health outcomes and be connected to existing programs to address their needs [16]. Leveraging SDoH data within EHRs can activate embedded tools like alerts and flags, guiding interventions like nutrition assistance based on hunger scores [17], community health worker referrals for those in disadvantaged neighborhoods [18], and creating high-risk patient panels for targeted care [19]. Such integration facilitates personalized care management and supports health equity through patient-centric technologies like self-scheduling apps [20] and digital navigators [21], fostering a learning health system that continuously adapts to emerging patient needs and outcomes.

**Figure 1.**
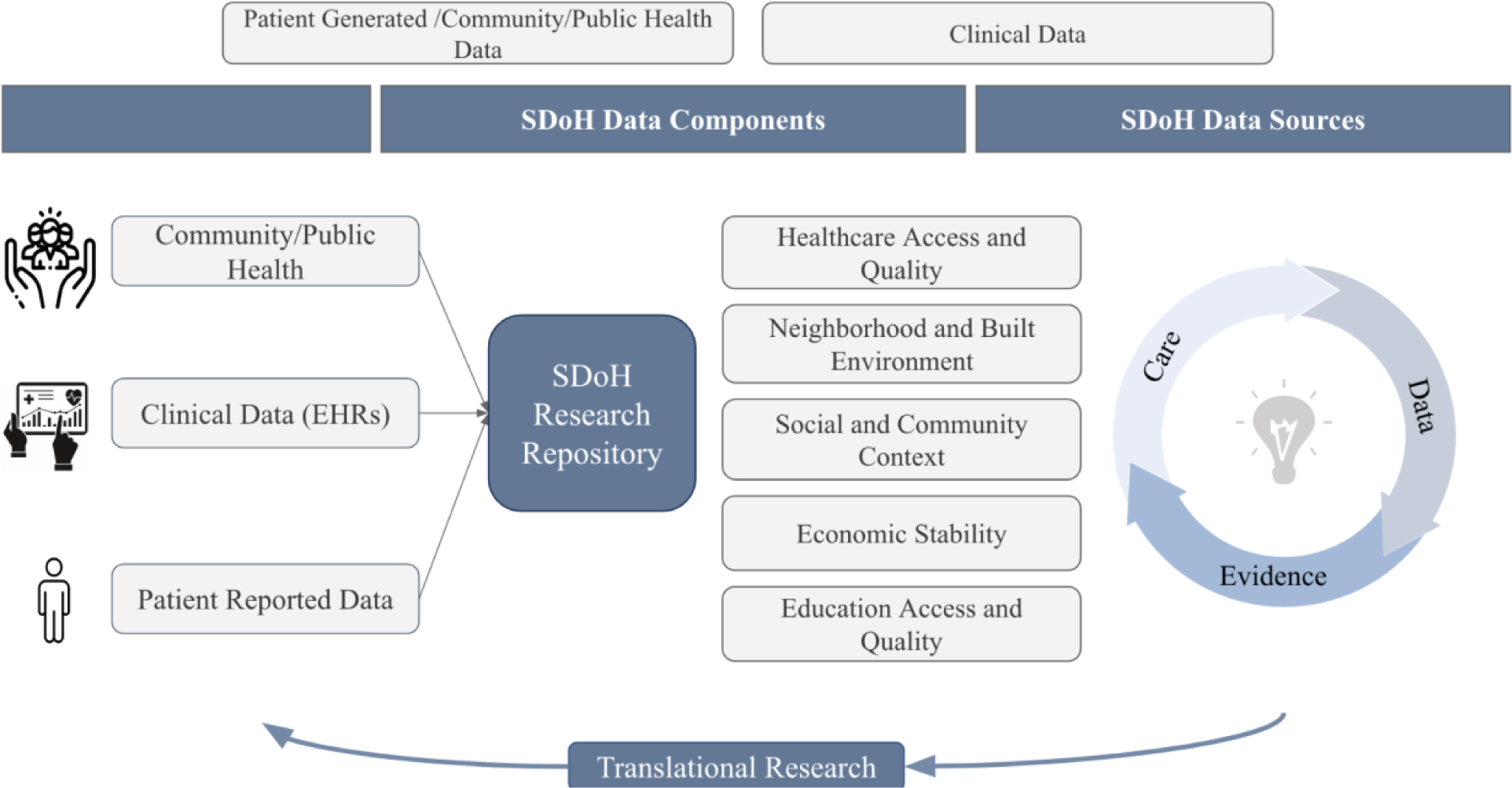
Data-to-Knowledge-to-Action Workflow for Translating SDoH into Clinical Care.

### Challenges and Barriers

Integrating SDoH data into EHRs is essential for enabling better decision-making and research to promote health equity, yet significant barriers exist [3,22]. Primarily, data on key factors like food insecurity, housing, transportation, social isolation, and financial strain are inconsistently and incompletely in structured data fields of the EHR [22,23].

Furthermore, healthcare professionals use validated screening tools, but of inconsistent types with different data standards, which limits interoperability across systems [24]. On many occasions, clinical and administrative workflows do not facilitate routine collection and updating of SDoH data over time. [25,26]

From a regulatory standpoint, confidentiality rules combined with patients’ mistrust of how private information is disclosed and shared prevent SDoH data from being shared for extended application [27]. A multi-pronged approach involving policy change, system redesign, and community engagement is imperative to fulfill the potential of SDoH [28].

Preliminary results demonstrated that important SDoH factors such as food insecurity, transportation barriers, and unstable housing were commonly discussed in clinical encounters across a range of specialties. However, these social needs were rarely codified in discrete structured fields. This finding aligns with prior literature suggesting substantial gaps in systematic SDoH data capture in EHR systems [29]. While NLP approaches may help unlock SDoH data trapped in free-text notes, a more robust data collection and integration framework is needed to optimize SDoH data capabilities in EHRs.

Once it is known that a patient is experiencing an SDoH and carries risks of associated poor health outcomes, e.g., 30-day readmission, it is critical for the care providers to connect patients to existing programs and services to mitigate the risk by addressing the disparity. However, it can be challenging to know what programs and services are available to the patient and whether the patient meets enrollment criteria. Clinical decision support systems and other digital health technologies could play a critical role in determining patient eligibility criteria and bringing service recommendations to care providers, e.g., case management or discharge planning teams [30].

While prior works have reviewed aspects of SDoH data collection, NLP approaches, and health impacts separately, a comprehensive overview integrating evidence across domains could further knowledge. This scoping review aimed to map the literature landscape surrounding SDoH-EHR integration to address several key questions:

1) *What standardized tools and workflows currently exist for structured SDoH data capture in EHRs?*
2) *How are external SDoH data sources linked to patient records to enable enriched contextual patient profiles?*
3) *What NLP solutions show promise for extracting unstructured SDoH insights from clinical notes at scale?*
4) *What impacts do harmonized SDoH data elements have on predicting health outcomes and targeting interventions?*

By systematically searching evidence related to these questions, this review identifies current best practices, remaining gaps, and future directions across the spectrum of SDoH data integration. The goal is to advance standardized frameworks for reliably collecting multidimensional social data within EHRs and translating derived knowledge to improve care delivery and health equity.

## Method

### Scoping Literature Review

To explore the current landscape of integrating SDoH data into EHRs, we conducted a scoping literature review. This methodological approach allows for a comprehensive overview of a broad field of study, and is particularly suitable for fields that have yet to be comprehensively reviewed or where the literature is large, diverse, and complex.

### Search Strategy

The literature search was conducted in PubMed on 2023 May 8th, a widely-recognized database for biomedical literature. We utilized MeSH (Medical Subject Headings) terms to refine our search, focusing on articles indexed with terms “Electronic Health Records” and “Social Determinants of Health.” This combination was chosen to specifically target studies that discuss the intersection of EHRs with SDoH (Search strategy see **Supplement Table 1**).

### Screening and Selection process

The inclusion process involved an initial title and abstract screening phase to identify papers related to SDoH capabilities and EHRs.

Selected papers were categorized into five non-mutually exclusive, key topics to structure our analysis (depicted in **Figure 2)**:

● **SDoH Screening Tools and Assessments**: Papers discussing various tools and methodologies for screening SDoH.
● **SDoH Data Collection and Documentation**: Studies focusing on how SDoH data is collected and documented within EHR systems.
● **Use of Natural Language Processing (NLP) for SDoH:** Research exploring the application of NLP techniques to identify and extract SDoH information from unstructured EHR data.
● **Associations between SDoH and Health Outcomes:** Papers examining the relationship between SDoH and various health outcomes.
● **SDoH Interventions:** Studies that evaluate the effectiveness of interventions aimed at addressing SDoH within healthcare settings.

**Figure 2.**
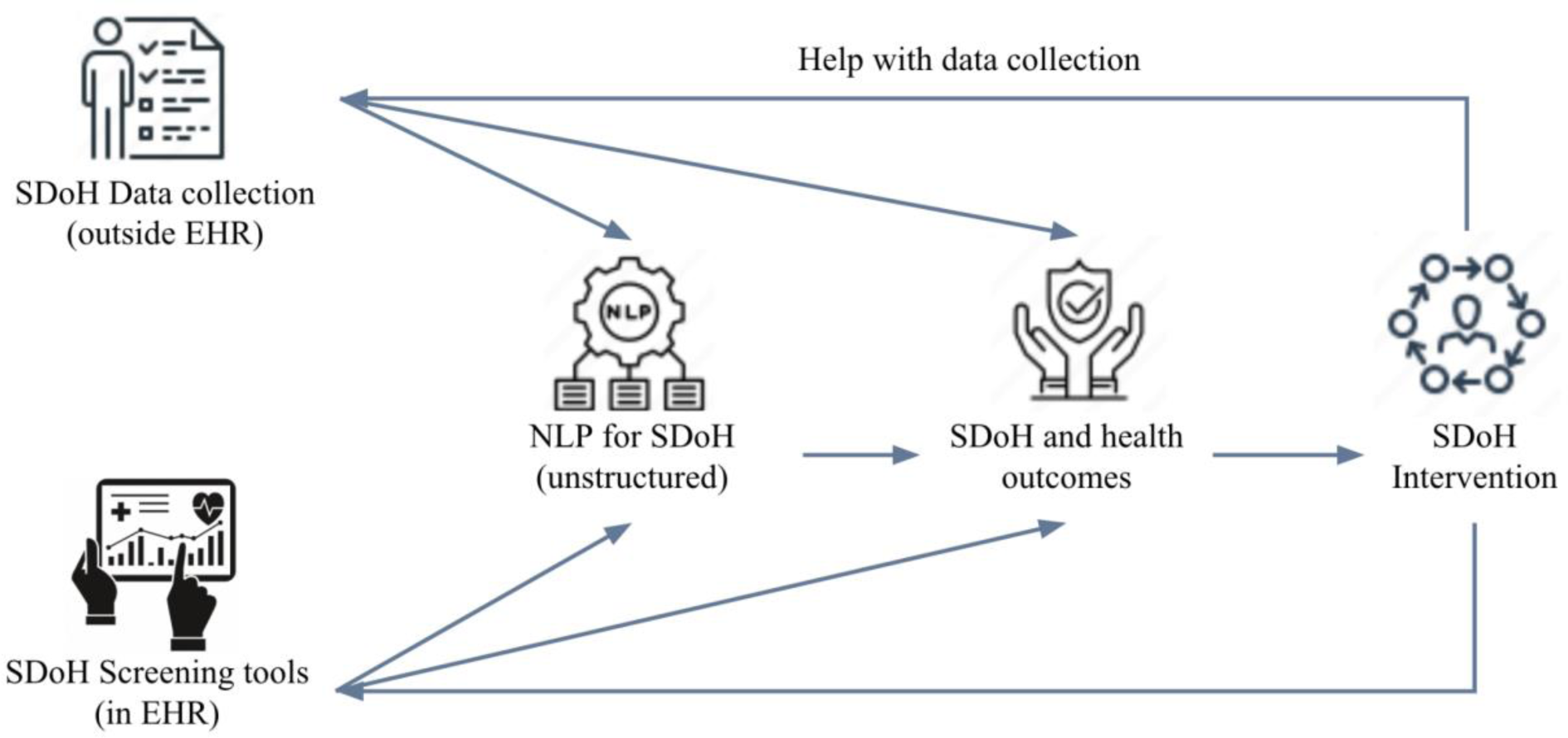
Five SDoH categories describing the data workflow from data capture efforts to interventions.

In phase two, junior authors were randomly assigned manuscripts for initial meta-data extraction. The full text screening involved detailed examination to confirm studies had substantive focus on the screening tools, data harmonization techniques, text analytics, associations, or interventions sub-topics. Additional irrelevant studies were excluded in this stage.

Phase three then consisted of evidence synthesis and conflict resolution by senior authors, who double-checked and verified the accuracy and completeness of extractions to enhance consistency. Through this additional quality assurance process, senior authors validated phase one and two results. The PRISMA flow diagram[31] was utilized to depict the screening process, detailing the numbers of identified, included and excluded studies across the systematic search and screening phases, along with reasons for exclusion.

The multi-stage screening and extraction process with independent categorization, full-text meta-data extraction, and consensus meetings helped embed quality checks aligning with scoping evidence best practices.

## Results

In this scoping review, we present the findings of SDoH in the EHR according to 5 domains of interest.

### Data Collection and synthesis

The initial PubMed query resulted in 685 articles. Articles were reviewed for inclusion based on their titles and abstracts resulting in 415 articles. Of these 415 articles, 324 articles included full text for qualitative synthesis. The reviewed articles were then classified according to SDoH in the EHR domains. The majority of articles focused on **SDoH and health outcomes, SDoH data collection and documentation** followed by **NLP for SDoH,** then **SDoH screening tools and assessments**. In the following sections, we reviewed the major themes and highlighted works for each of the five SDoH in the EHR domains (see **Figure 3**, percentage see **Supplement Table 2**).

**Figure 3.**
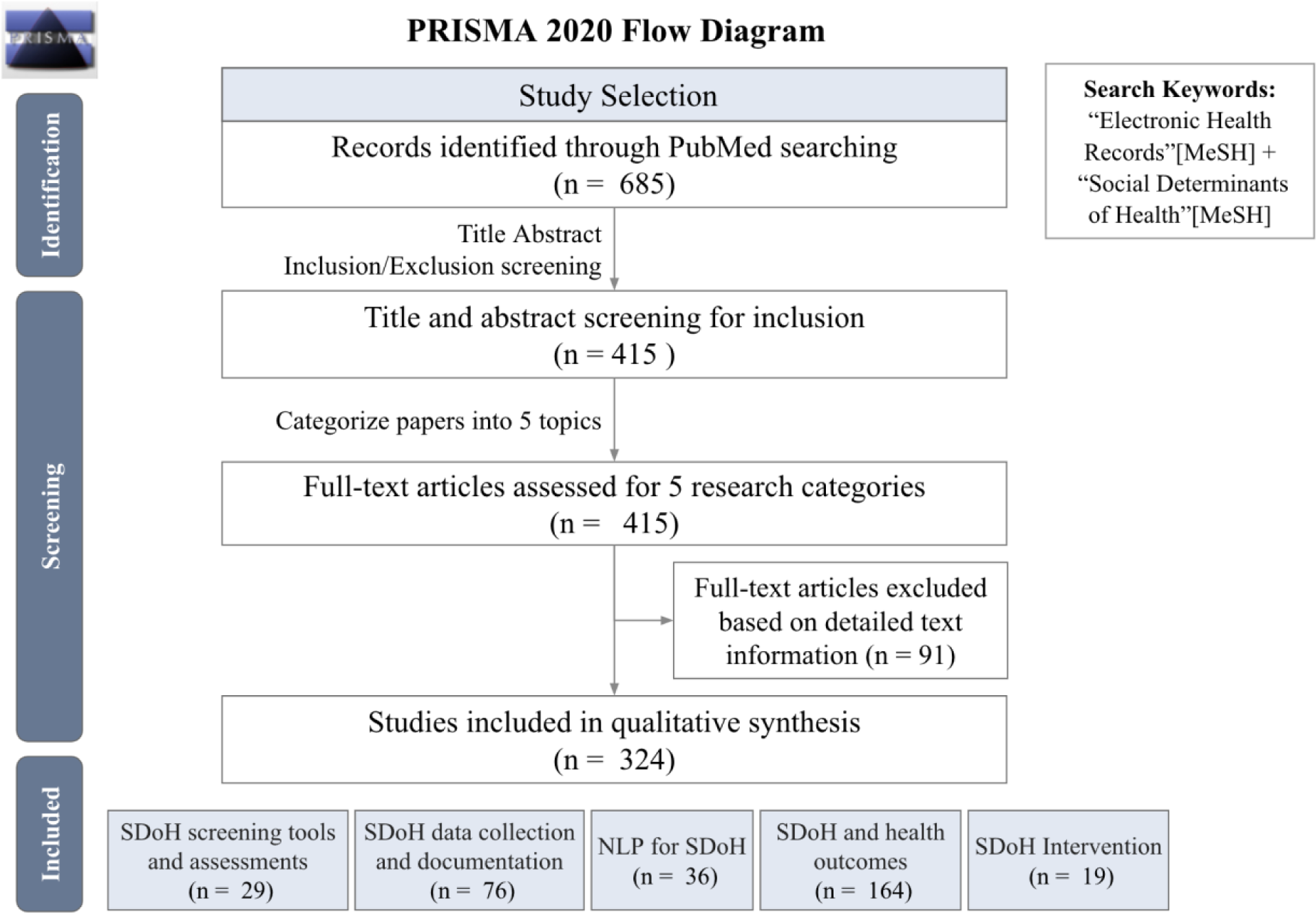
PRISMA 2020 Flow Diagram.

Aligned with PRISMA guidelines (Tricco [31], the screening process involved an initial title/abstract review phase led by author C.L. to categorize papers into one or more of the 5 topics. Targeted meta-data extraction was performed by assigned reviewers as follows: Screening Assessments (R.Y.), SDoHData Collection (C.L.), NLP Approaches (C.L.), and Interventions (X.M.). The SDoH and Outcomes papers were randomly assigned to the broader reviewer pool (C.L., R.Y., S.H., D.L.M., U.V., H.K.D.) for meta-data extraction. Additional irrelevant studies were excluded in this second phase (see **Table 2** for details). The full-text meta-data extraction phase allowed confirmation of accurate categorization and extraction, with discrepancies resolved through consensus meetings. Evidence synthesis leads included: C.L. for SDoH Screening tools and SDoH and health outcomes, D.L.M for SDoH Data collection, NLP for SDoH), X.M. for SDoH Interventions, overseen by senior authors M.J.M. and D.L.M.

**Table 1.** WHO-SDoH Data Components.

| SDoH concept | SDoH domains |
| --- | --- |
| Economic Stability | Income, employment, financial resources, and basic needs |
| Neighborhood and Built Environment | Neighborhood (zip code), transportation, food access, environmental conditions |
| Health and Health Care | Insurance status (type, coverage, payer, provider), behavioral and mental health |
| Education | Attainment, level, language |
| Social and Community Context | Race/ethnicity, connections, marital status |

**Table 2.** Title Abstract Inclusion/Exclusion Criteria.

|  |
| --- |
| **Inclusion Criteria:**<br>• Original empirical research studies, including case studies, technical reports, and database studies<br>• Studies centrally focused on integrating/analyzing SDoH data elements in electronic health records systems<br>• Addressed one or more of following defined five domains<br>• Provided details on methods, data sources, outcomes, targets related to SDoH factors<br>• Published in English language |
| **Exclusion Criteria:**<br>• Review papers, editorials, opinion pieces, conceptual frameworks<br>• Informatic models/approaches without tie to real EHR/health database<br>• Purely qualitative studies lacking SDoH-EHR data focus<br>• Biomedical studies incorporating socioeconomic variables, but lacking central SDoH-EHR focus<br>• Lacking details on techniques/sources for ingesting/analyzing SDoH data<br>• Study protocols without substantive findings section |

**Table 3.**
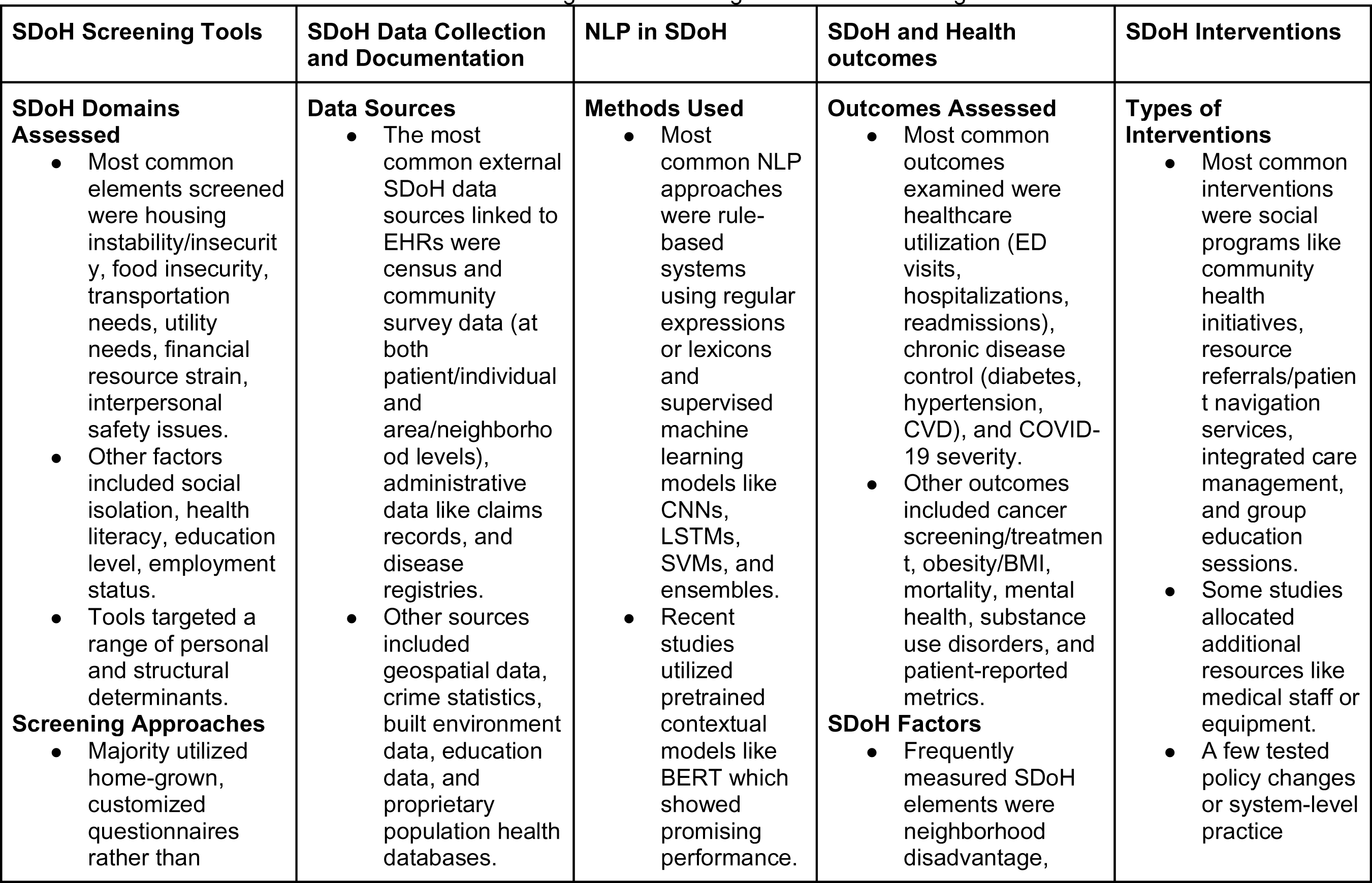
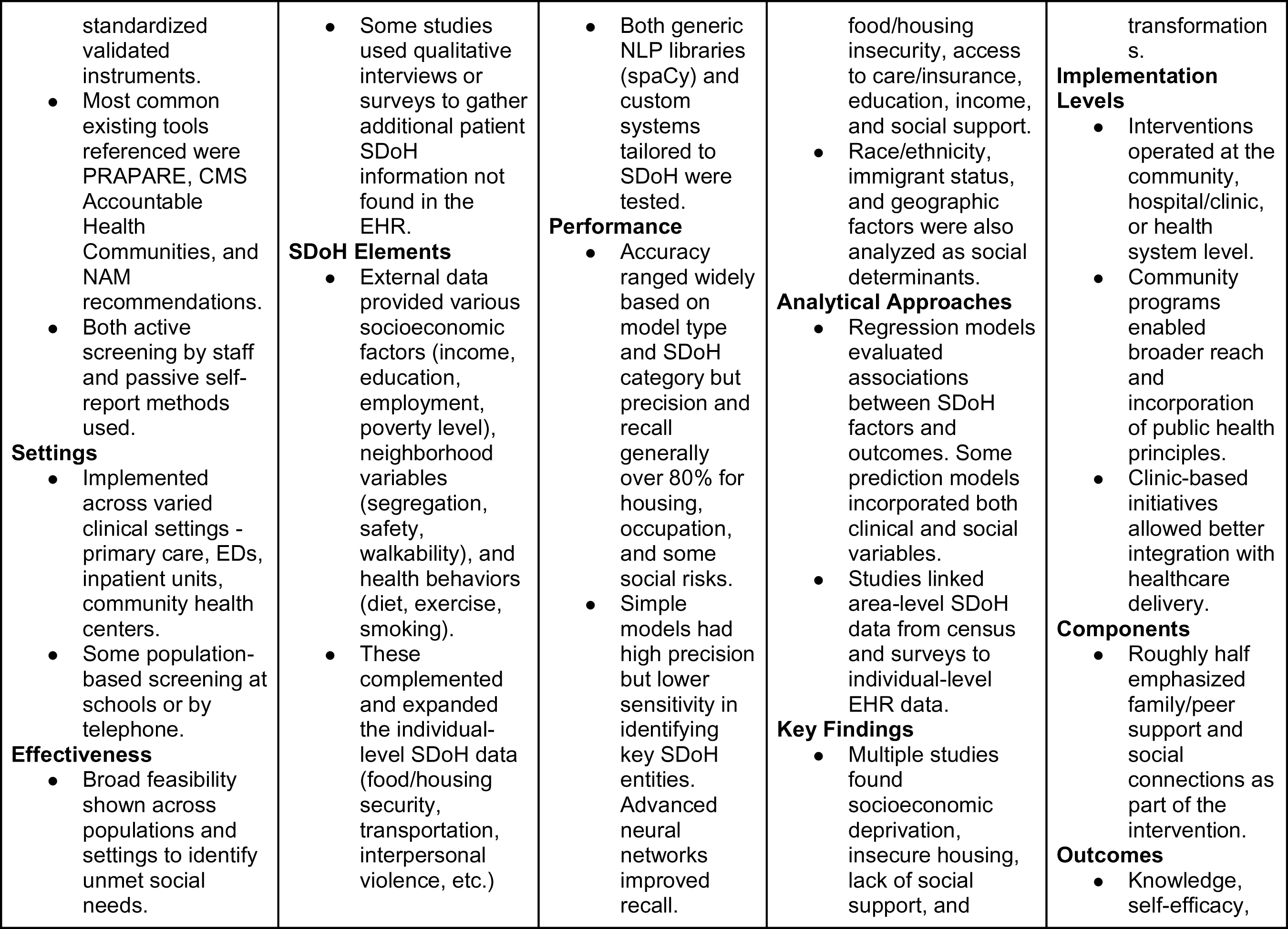
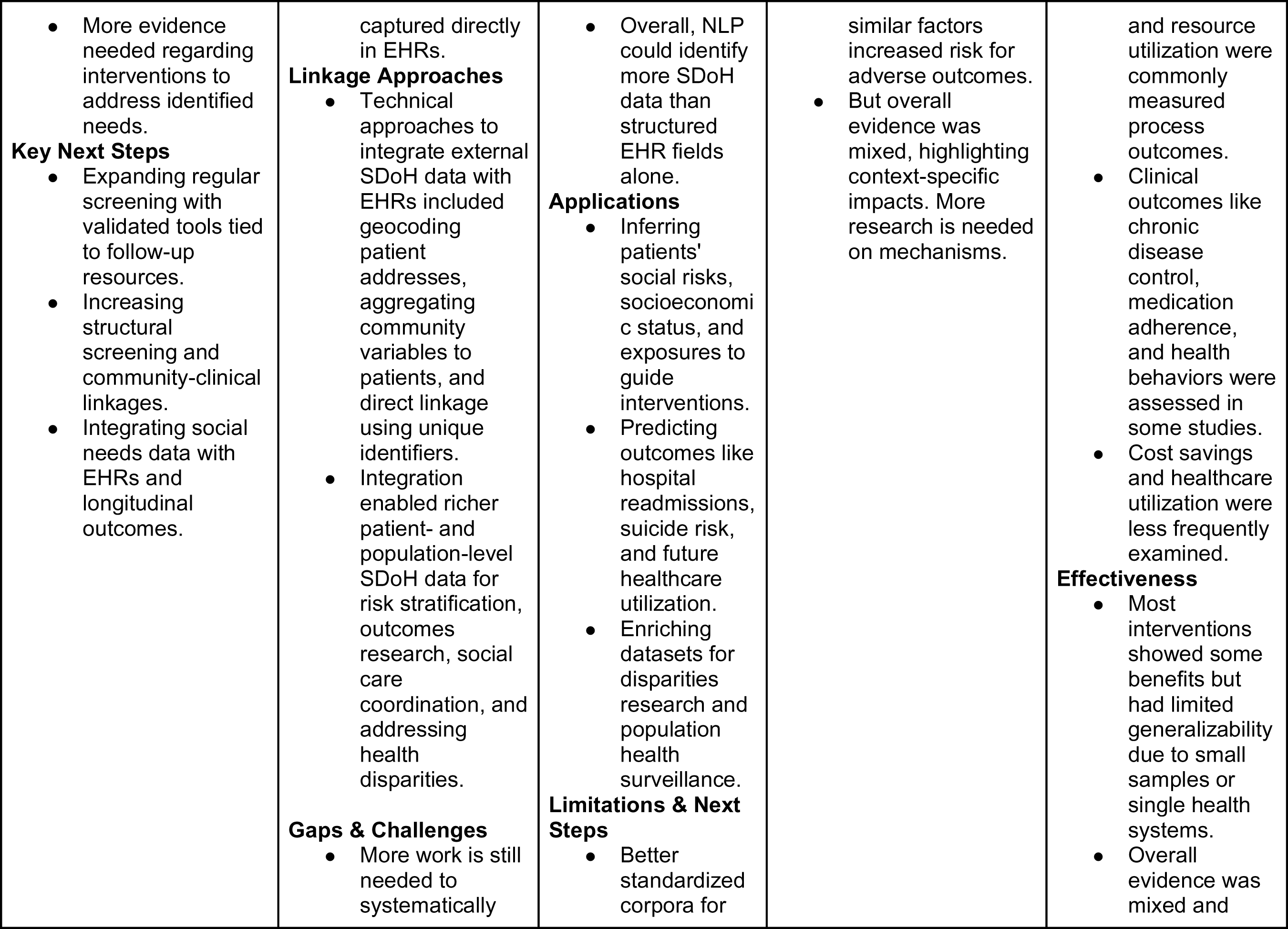
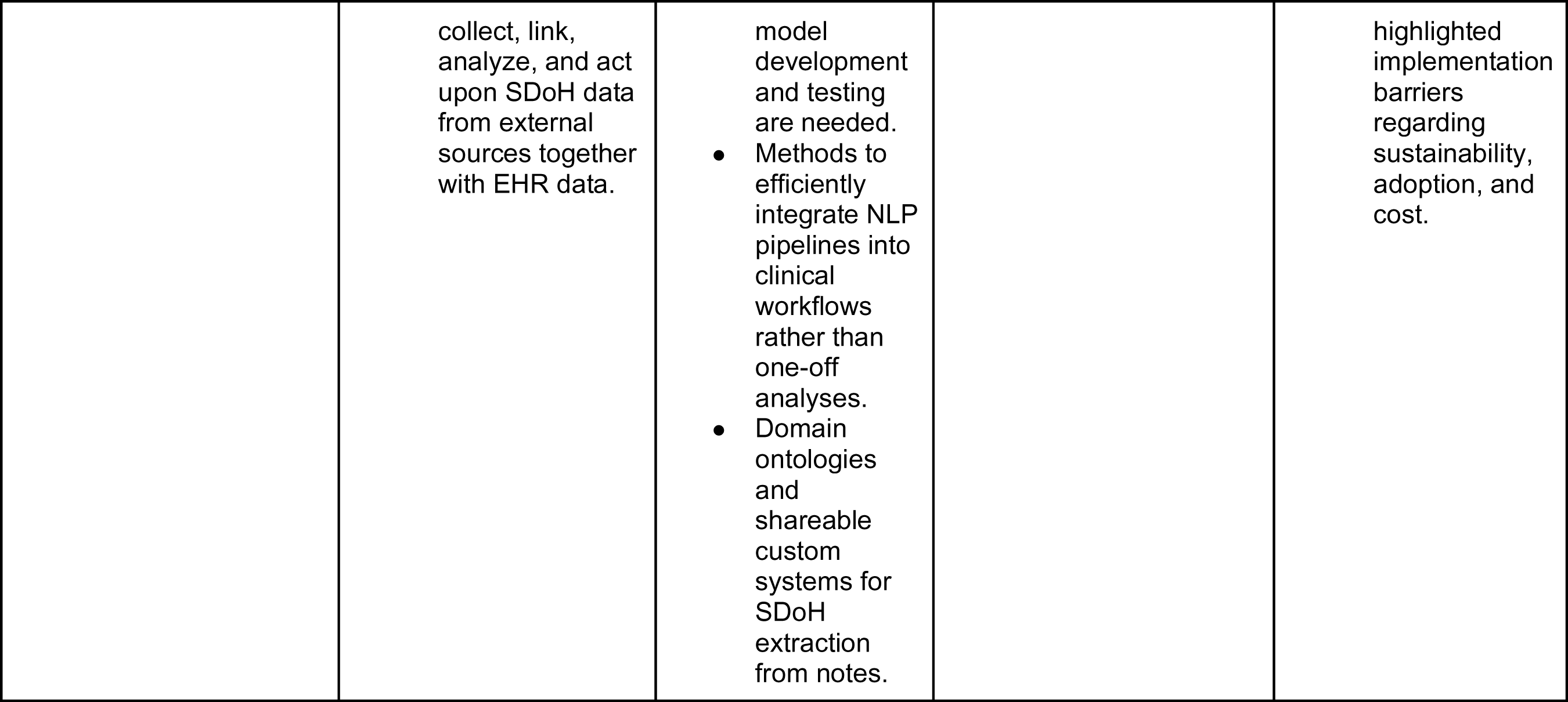
SDoH-driven Translational Research: Deriving and Translating Actionable Knowledge into Clinical Care.

| SDoH Screening Tools | SDoH Data Collection and Documentation | NLP in SDoH | SDoH and Health outcomes | SDoH Interventions |
| --- | --- | --- | --- | --- |
| **SDoH Domains Assessed**<br>• Most common elements screened were housing instability/insecurity, food insecurity, transportation needs, utility needs, financial resource strain, interpersonal safety issues.<br>• Other factors included social isolation, health literacy, education level, employment status.<br>• Tools targeted a range of personal and structural determinants.<br>**Screening Approaches**<br>• Majority utilized home-grown, customized questionnaires rather than | **Data Sources**<br>• The most common external SDoH data sources linked to EHRs were census and community survey data (at both patient/individual and area/neighborhood levels), administrative data like claims records, and disease registries.<br>• Other sources included geospatial data, crime statistics, built environment data, education data, and proprietary population health databases. | **Methods Used**<br>• Most common NLP approaches were rule-based systems using regular expressions or lexicons and supervised machine learning models like CNNs, LSTMs, SVMs, and ensembles.<br>• Recent studies utilized pretrained contextual models like BERT which showed promising performance. | **Outcomes Assessed**<br>• Most common outcomes examined were healthcare utilization (ED visits, hospitalizations, readmissions), chronic disease control (diabetes, hypertension, CVD), and COVID-19 severity.<br>• Other outcomes included cancer screening/treatment, obesity/BMI, mortality, mental health, substance use disorders, and patient-reported metrics.<br>**SDoH Factors**<br>• Frequently measured SDoH elements were neighborhood disadvantage, | **Types of Interventions**<br>• Most common interventions were social programs like community health initiatives, resource referrals/patient navigation services, integrated care management, and group education sessions.<br>• Some studies allocated additional resources like medical staff or equipment.<br>• A few tested policy changes or system-level practice |
| standardized validated instruments.<br>• Most common existing tools referenced were PRAPARE, CMS Accountable Health Communities, and NAM recommendations.<br>• Both active screening by staff and passive self-report methods used.<br>**Settings**<br>• Implemented across varied clinical settings - primary care, EDs, inpatient units, community health centers.<br>• Some population-based screening at schools or by telephone.<br>**Effectiveness**<br>• Broad feasibility shown across populations and settings to identify unmet social needs. | • Some studies used qualitative interviews or surveys to gather additional patient SDoH information not found in the EHR.<br>**SDoH Elements**<br>• External data provided various socioeconomic factors (income, education, employment, poverty level), neighborhood variables (segregation, safety, walkability), and health behaviors (diet, exercise, smoking).<br>• These complemented and expanded the individual-level SDoH data (food/housing security, transportation, interpersonal violence, etc.) | • Both generic NLP libraries (spaCy) and custom systems tailored to SDoH were tested.<br>**Performance**<br>• Accuracy ranged widely based on model type and SDoH category but precision and recall generally over 80% for housing, occupation, and some social risks.<br>• Simple models had high precision but lower sensitivity in identifying key SDoH entities. Advanced neural networks improved recall. | food/housing insecurity, access to care/insurance, education, income, and social support.<br>• Race/ethnicity, immigrant status, and geographic factors were also analyzed as social determinants.<br>**Analytical Approaches**<br>• Regression models evaluated associations between SDoH factors and outcomes. Some prediction models incorporated both clinical and social variables.<br>• Studies linked area-level SDoH data from census and surveys to individual-level EHR data.<br>**Key Findings**<br>• Multiple studies found socioeconomic deprivation, insecure housing, lack of social support, and | transformations.<br>**Implementation Levels**<br>• Interventions operated at the community, hospital/clinic, or health system level.<br>• Community programs enabled broader reach and incorporation of public health principles.<br>• Clinic-based initiatives allowed better integration with healthcare delivery.<br>**Components**<br>• Roughly half emphasized family/peer support and social connections as part of the intervention.<br>**Outcomes**<br>• Knowledge, self-efficacy, |
| • More evidence needed regarding interventions to address identified needs.<br>**Key Next Steps**<br>• Expanding regular screening with validated tools tied to follow-up resources.<br>• Increasing structural screening and community-clinical linkages.<br>• Integrating social needs data with EHRs and longitudinal outcomes. | captured directly in EHRs.<br>**Linkage Approaches**<br>• Technical approaches to integrate external SDoH data with EHRs included geocoding patient addresses, aggregating community variables to patients, and direct linkage using unique identifiers.<br>• Integration enabled richer patient- and population-level SDoH data for risk stratification, outcomes research, social care coordination, and addressing health disparities.<br>**Gaps & Challenges**<br>• More work is still needed to systematically | • Overall, NLP could identify more SDoH data than structured EHR fields alone.<br>**Applications**<br>• Inferring patients' social risks, socioeconomic status, and exposures to guide interventions.<br>• Predicting outcomes like hospital readmissions, suicide risk, and future healthcare utilization.<br>• Enriching datasets for disparities research and population health surveillance.<br>**Limitations & Next Steps**<br>• Better standardized corpora for | similar factors increased risk for adverse outcomes.<br>• But overall evidence was mixed, highlighting context-specific impacts. More research is needed on mechanisms. | and resource utilization were commonly measured process outcomes.<br>• Clinical outcomes like chronic disease control, medication adherence, and health behaviors were assessed in some studies.<br>• Cost savings and healthcare utilization were less frequently examined.<br>**Effectiveness**<br>• Most interventions showed some benefits but had limited generalizability due to small samples or single health systems.<br>• Overall evidence was mixed and |
|  | collect, link, analyze, and act upon SDoH data from external sources together with EHR data. | model development and testing are needed.<br>• Methods to efficiently integrate NLP pipelines into clinical workflows rather than one-off analyses.<br>• Domain ontologies and shareable custom systems for SDoH extraction from notes. |  | highlighted implementation barriers regarding sustainability, adoption, and cost. |

### SDoH Screening Tools

We included 29 papers (details see Supplement Table 3) incorporating SDoH Screening tools into EHRs in our review.The majority of the studies utilized home-grown tools for screening SDoH. Some studies [29,32–34] developed their own questionnaires and screening sets, reflecting a trend towards customized tools tailored to specific healthcare settings or populations. Vendor-specific tools, like the two-item screening tool [35] integrated into Epic SDoH Wheel, were less common, but still present. The screened determinants varied, but commonly included factors included housing, food insecurity, transportation, and mental health indicators like stress and depression.

Studies targeted a diverse range of populations. For example, children were the focus in some studies [32,36], while adults were the primary subjects in studies [33,37]. Various healthcare settings were represented, from primary care clinics [38,39] to emergency departments [40], as well as school-based clinics [34] indicating the widespread recognition of the importance of SDoH in different medical environments, underscoring the growing acknowledgment of SDoH’s relevance across the healthcare spectrum.

Active screening methods, where healthcare providers proactively administered questionnaires or interviews, were predominant (n = 28) [29,33]. Passive methods like the analysis of EHR data [41] were less common, but they represented an emerging trend. However, the utilization of EHR data for passive screening suggests an emerging trend that could streamline the process in the future. While many studies focused on personal health determinants (n = 19), others also assessed structural determinants like housing quality and social networks [32,39,42]. Not many studies (n = 8), looked at both personal and structural determinants.

### Challenges and Opportunities

The reviewed studies collectively highlight the challenges in standardizing SDoH screening across various contexts but also point towards the potential benefits of such screenings to patient care. The diversity in approaches, as seen in studies [29,32–34], reflects the complexity of addressing SDoH in clinical practice. However, it also demonstrates a concerted effort towards more comprehensive patient care. The prevalence of home-grown tools [32,34,39,43] indicates a trend towards customization, tailored to specific patient populations and healthcare settings. This is likely due to the unique needs and circumstances of different patient demographics. The variability in tools and approaches (e.g., the number of questions in studies [29,44], and the use of paper-based vs. EHR-based tools [33,38] highlight the challenges in standardizing SDH screening. This variability could impact the comparability of data and the scalability of successful approaches. Despite the challenges, the focus on SDoH screening illustrates a shift towards patient care. Recognizing and addressing social and behavioral factors [34,45] can lead to more effective healthcare interventions and better health outcomes.

### SDoH Data Collection and Documentation

In our review, we identified 76 articles (details see Supplement Table 4) describing SDoH data collection and documentation practices. Studies focused on engagement with populations and leveraged a variety of technologies to support collection and documentation processes. Few studies (n = 9) included interviews of patients and clinicians, engagement with communities, focus groups, town hall meetings and the like [46–54]. Other studies (n = 7) collected SDoH information using paper-based entry, iPads/tablets, patient or clinician-facing web portals and other web-based toolkits and forms [55–61].

Several studies made use of publicly available, external data resources to infer structural SDoH information for a given population. The most common external SDoH data sources linked to EHRs were US census and community survey data (at both patient/individual and area/neighborhood levels), administrative data/claims records, and disease registries. Commonly linked community surveys and systems include the Behavioral Risk Factor Surveillance System (BRFSS), the National Health and Nutrition Examination Survey (NHANES), the National Health Interview Survey (NHIS) [62], the National Institutes of Health PROMIS® (Patient-Reported Outcomes Measurement Information System)[63], the National Survey of Children’s Health (NSCH)[64], the Center for Disease Control Youth Risk Behavior Surveillance System (YRBSS) [65], the Center for Medicare and Medicaid Services (CMS) Accountable Health Communities’ Health-Related Social Needs Screening Tool [66], the National Center for Education Statistics, the Uniform Crime Reports, and the American Community Survey [67–70].

Longitudinal study data included the National Longitudinal Study of Adolescent to Adult Health (Add Health) [71]. Other administrative data sources included the HCUP Nationwide Readmissions Database [72], claims data [73], and Medicaid data warehouse [74]. Few studies describe use of disease-specific registries e.g. cancer registries such as SEER-CMS, SEER-Medicare, SEER-Medicaid [75].

Several studies (n = 8) describe methods for inferring structural SDoH using geocoding of patient addresses and linking to public census tract data [60,76–78] to integrate information related to neighborhood and community-level characteristics (e.g., SES, crime incidence, health facility locations) [79] and neighborhood factors (e.g., poverty level, education, employment status, etc.) [68,80,81].

External data provided various socioeconomic factors (income, education, employment, poverty level, air quality), neighborhood variables (segregation, safety, walkability), and health behaviors (diet, exercise, smoking) [62,76,82–86]. These complemented and expanded the individual-level SDoH data (food/housing security, transportation, interpersonal violence, etc.) captured directly in EHRs [25,26,28,46,73,82,84,87–93].

Few studies (n = 3) aimed to integrate EHR, genomic, and public health data to study the intersection of lifestyle, genetics, and environmental influences [94–96]

### Challenges and Opportunities

Although these works highlight the potential for study of personal and structural SDoH, there is considerable effort for systematically collecting, linking, and analyzing SDoH data from external sources together with EHR data at the community, state, and national levels [84,97,98]. Few studies have incorporated the use of common data models for improving standardization and interoperability of collected SDoH data [99–101]. Furthermore, few studies demonstrated how this information could be leveraged to connect individuals with SDoH risk factors to social programs.

### NLP in SDoH

In our review, we identified 36 articles (details see Supplement Table 5). describing NLP methods for powering SDoH studies. Many SDoH elements are locked within clinical free-text notes requiring NLP for identification, encoding, and extraction to standardized terminologies such as SNOMED-CT or representations [102]. Few studies focused on lexicon development using methods such as lexical associations, word embeddings, term similarity, and query expansion. Lexicons and regular expressions have been demonstrated to extract SDoH and psychosocial risk factors [103–105], learn distinct social risk factors with mappings them to standard vocabularies and code sets including ICD-9/10, ICD Z codes, **U**nified **M**edical **L**anguage **S**ystem (UMLS), and SNOMED-CT. Most articles (n = 15) describe rule-based approaches using regular expressions and/or hybrid machine learning methods leveraging platforms. Some articles (n = 5) highlighted well-known rule-based toolkits and platforms adapted with lexicons and regular expressions for SDoH extraction including Moonstone, **Easy C**linical **I**nformation **E**xtraction **S**ystem (EasyCIE), **M**edical **T**ext **E**xtraction, **R**easoning and **M**apping **S**ystem (MTERMS), **Q**ueriable **P**atient **I**nference **D**ossier (QPID), and **Cl**inical **Eve**nt **R**ecognizer (CLEVER) [19,106–109]. Other articles (n = 7) describe rule-based systems paired with traditional machine learning approaches i.e., an ensemble, particularly using NLP systems such as **G**eneral **A**rchitecture for **T**ext **E**ngineering (GATE), **C**linical **L**anguage **A**nnotation, **M**odeling, and **P**rocessing Toolkit (CLAMP), **E**xtr**a**ct **S**DOH from **E**HRs (EASE), Yale **c**linical **T**ext **A**nalysis and **K**nowledge **E**xtraction **S**ystem (cTAKES), **Re**lative **Hou**sing **S**tability in **E**lectronic **D**ocumentation (ReHouSED), and toolkits such as spaCy and medspaCy in conjunction with conditional random fields and support vector machines [110–113]. In contrast, several investigators have leveraged open-source NLP toolkits like spaCy and medspaCy without supervised learners to extract SDoH variables [114–116]. Other studies (n = 19) have solely leveraged traditional supervised and unsupervised learning techniques, support vector machines (SVM), logistic regression (LR), Naïve Bayes, Adaboost, Random Forest, XGBoost, Bio-ClinicalBERT, Latent Dirichlet Allocation (LDA), and bidirectional Long Short-Term Memory (BI-LSTM) [16,117–121] to extract and standardize social and behavioral determinants of health (SBDoH), e.g., alcohol abuse, drug use, sexual orientation, homelessness,substance use, sexual history, HIV status, drug use, housing status, transportation needs, housing insecurity, food insecurity, financial insecurity, employment/income insecurity, insurance insecurity, and poor social support. In more recent years, studies (n = 9) have focused on the training and tuning of deep learning approaches, primarily transformer-based approaches i.e., Bidirectional Encoder Representations from Transformers (BERT), RoBERTa, BioClinical-BERT models for extracting SBDoHs including relationship status, social status, family history, employment status, race/ethnicity, gender, social history, sexual orientation, diet, alcohol, smoking housing insecurity, unemployment, social isolation, and illicit drug use—from clinical notes, PubMed, among other specialised texts e.g., LitCOVID [122–131]. The frequency of papers for SDoH extraction NLP algorithms within EHR systems, highlighting the combinations and intersections of utilized methodologies can be found in **Figure 4**.

**Figure 4.**
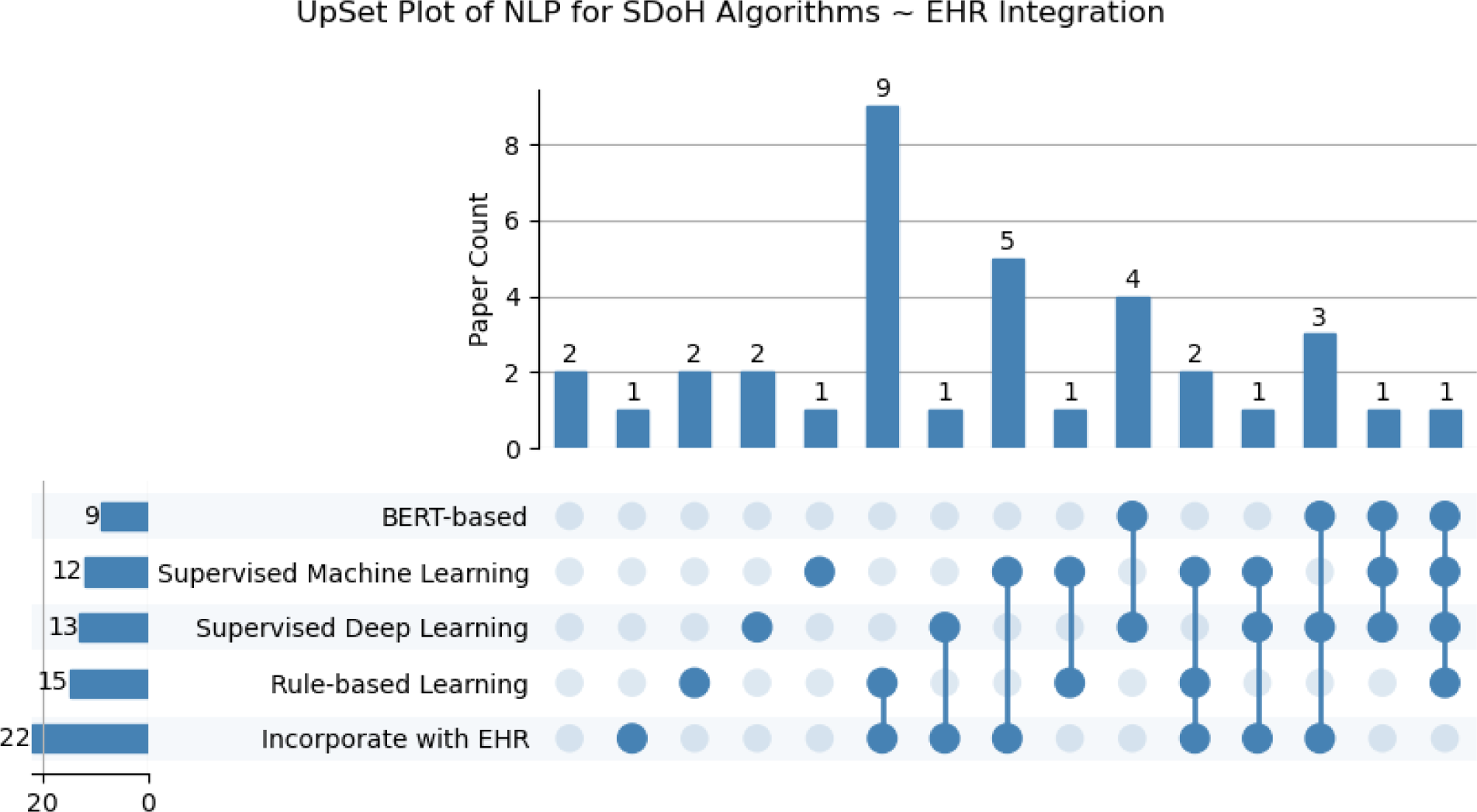
UpSet Plot of NLP for SDoH Algorithms ~EHR Integration. *Outlining the distribution of papers in which each approach/method individually and in combination was described in the study. *Supervised Machine Learning includes traditional machine learning methods (Naive Bayes, SVM, logistic regression, random forest, etc), excluding neural networks and pre-trained approaches.

### Challenges and Opportunities

Although these works highlight the potential for extracting SDoH from texts, several challenges remain. Few studies focused on lexicon development, make use of standard terminologies for encoding SDoH data, and explore deep extraction and representation of SDoH attributes and relationships. Also, many studies focus on extraction and encoding SDoH data from a single site and fail to assess the portability of methods to new textual data sources beyond clinical notes and PubMed articles such as digital technologies and chatbots. The introduction of shared datasets like the Social History Annotated Corpus (SHAC) dataset is an important step towards demonstrating generalizability of NLP-powered, SDoH extraction systems. Emerging generative models may also improve upon the state-of-the-art demonstrated by common shared task datasets.

### SDoH and Health Outcomes

In our review, we identified 164 articles (details see Supplement Table 6). describing SDoH and health outcomes. SDoH and health outcome studies examined a wide variety of health-related events and outcomes in relation to SDoH factors. A predominant focus was on infectious disease outcomes, with 16 studies examining drivers of COVID-19 hospitalization, mortality, treatment disparities and differences in positivity rates across social groups [132–135]. Another major category included healthcare utilization metrics like preventable hospital readmissions (n = 11) [136–145], ED reliance (n = 16) [146–168], and telehealth adoption [169]. Beyond infectious outcomes and healthcare utilization, studies also assessed chronic disease control across conditions like diabetes (n = 11) [170–180], hypertension [173,181,182], kidney disease [183,184], and obesity (n = 7) [146,175,185–189], along with risk factors like elevated blood pressure and cardiovascular events. Some studies focused on cancer (n = 11) screening, diagnoses, treatment disparities and survival outcomes [167,190–199], while others addressed mental health (n = 6) indicators [146–151] ranging from dementia incidence [200] to suicide (n = 2) risk factors [201,202]. Additional outcomes evaluated included maternal morbidity (n = 2) [203,204], pediatric health metrics ranging from vaccine completion rates to epilepsy-related consequences.

The most common quality measures reported were standardized condition control thresholds like HbA1c levels for diabetes control [173,205], blood pressure (n = 5) levels for hypertension control, and established cancer staging guidelines [191]. Some studies used validated risk prediction models for outcomes like hospital readmissions (n = 5), suicide risk [201] or mortality (n = 6). Beyond clinical indicators, several studies incorporated validated SDoH indexes like the Area Deprivation Index [206,207], Social Deprivation Index [208] and CDC’s Social Vulnerability Index[209]. In terms of analysis approaches, common methods included multivariate regression models like logistic regression (n = 13) [159,179,180,192,199,210–217] and Cox proportional hazards models (n = 3) [138,196,212] to assess adjusted outcome associations with SDoH factors. Other advanced techniques leveraged included machine learning algorithms [215], geospatial analysis for clustering [218], and phenome-wide association studies [150] in select studies.

### SDoH Interventions

A total of 19 papers (details see Supplement Table 7). were found that collected supplementary SDoH data to support population health intervention initiatives targeting hospitals/clinics (n = 10) or communities (including primary care, n = 9) at the meso (institution) level. Two articles discussed policy potential and proposed policy reform at the macro (system) level [219,220]. The majority of the selected research (n = 16) focused on implementing a social and healthcare supportive program to address the social needs of the target population. Interventions were implemented in various settings for hospital-based initiatives, including post hospital discharge [221], the emergency department [222], and clinics specializing in different medical disciplines [223–227]. On the other hand, community-based initiatives concentrated mainly on integrating interventions into primary care services [228–231]. The social and healthcare supportive programs included a range of initiatives designed towards improving community health. These initiatives encompassed the introduction of new healthcare programs [232,233], health education and coaching [225,232,234], the strengthening of medical-legal partnerships [228], the enhancement of integrated care planning [229,231,235] and the improvement of patient navigation [222,236,237]. Only a few papers have examined the potentials of incorporating the family or social support element into their intervention design [223,232,234]. Meanwhile, 4 studies investigated the potential for enhancing resource allocation through surveying outcomes. The improvement objectives encompassed the allocation of staff and equipment [221], the enhancement of patient navigation [224], and the transformation of health service practices [226,238].

Health outcomes, such as improvements in health metrics, reductions in disease incidence, changes in vital signs, and quality of life, are commonly used as measures to determine the feasibility of initiating an intervention (n = 10). Several studies have also assessed social SDoH in relation to patient satisfaction and acceptability [230,235,237,238]. Since most programs were new efforts, it was not possible to determine the effectiveness of the intervention in the short term, nor could the potential generalizability be assessed.

## Discussion

This scoping review set out to map SDoH-EHR integration literature across five key domains: structured data capture tools, external data linkage approaches, NLP-based extraction techniques, and applications for outcomes analysis, and health care interventions. Our discussion synthesized major themes and collective gaps within each sphere. Regarding our first aim, predominant tailored screening instruments enable assessment but standardization barriers persist. For the second objective, enriching patient profiles via claims and census linkage shows promise but systematic consolidation is lacking. On research question three, rule-based systems boast precision while neural networks improve unstructured element recognition—yet reproducibility hurdles remain. Finally, concerning predicting outcomes and targeting programs, consistent risk evidence conflicts with implementation uncertainty. Across objectives, we fulfill vital scoping aims in benchmarking maturation levels and specifying needs for advancing frameworks. Our cross-domain perspective illuminates such interdependencies requiring a “full-stack” approach. Thereby, we strengthen capacity for wisely collecting and translating multifaceted social data to guide health equity solutions.

## Key Findings by Theme

### SDoH Screening Tools

The studies implemented a variety of screening tools to assess patients’ social determinants of health (SDoH) across diverse healthcare settings. The most prevalent SDoH domains screened included housing instability/insecurity, food insecurity, transportation and utility service needs, interpersonal safety/violence, financial resource strain, social isolation, health literacy, and education level. Notably, the majority utilized home-grown screening instruments developed within their health systems rather than relying on standardized validated tools. The most commonly referenced standardized tools were PRAPARE, the National Academy of Medicine’s recommendations, and the Center for Medicare and Medicaid Services’ Accountable Health Communities screening tool. Researchers tested these tools across primary care clinics including family medicine, internal medicine, pediatrics, obstetrics/gynecology, and community health centers. Some studies also examined screening acceptance in emergency departments and inpatient units. While most relied on active screening conducted by providers during visits, several studies explored passive methods like paper questionnaires or electronic tablets. The instruments largely focused on assessing individual-level SDoH rather than community or structural factors. However, a minority did attempt to capture measures of both. Across diverse populations and implementation strategies, researchers found SDoH screening feasible to conduct and able to successfully identify unmet social needs amongst patients. Still further research remains warranted, particularly regarding optimal referral systems and interventions to address identified needs.

### SDoH Data Collection and Documentation

The current body of research demonstrates that numerous external data sources have been linked with electronic health records (EHRs) to enhance the capture of social determinants of health (SDoH). The most common linkages have connected individual and area-level census metrics, community surveys, administrative claims data, and disease registries to EHRs. Other integrated sources encompassing geospatial data, crime statistics, built environment factors, education data, and proprietary population health platforms offer additional context. Qualitative interviews and surveys have also supplemented patient SDoH insights not routinely documented in the course of clinical care [239]. These external data elements provide critical information surrounding socioeconomic position, neighborhood landscape, and health behaviors. By uniting individual and ecological variables, researchers can assemble more holistic patient profiles to advance risk stratification, outcomes studies, care coordination, and health equity initiatives [27,240].

Technical approaches underlying external SDoH data integration with EHRs have harnessed geocoding of addresses, aggregation of community measures, and linkage based on unique identifiers. While progress has occurred, further research must promote systematic collection, analysis, and application of integrated data sources in practice. Key steps for the field center on implementing reliable linkage mechanisms for disparate datasets and embedding multidimensional patient social profiles within clinical decision tools and workflows [26,241,242]. Only through purposeful integration and translation efforts can external SDoH data fully support identification of at-risk populations, patient-centered risk assessments, and targeted community-clinical interventions.

### NLP in SDoH

A range of natural language processing (NLP) approaches have been leveraged to identify critical social determinants from unstructured clinical notes. Common methods include rule-based systems using expert-curated lexicons and regular expressions as well as supervised machine learning models like convolutional neural networks and recurrent neural networks. More advanced studies have also employed contextual embedding models such as BERT and achieved promising performance. Both generic NLP software libraries and custom systems tailored to the social and behavioral health domains have been implemented. Reported accuracy metrics vary substantially by model type and target social determinant, but precision and recall generally exceed 80% for housing insecurity, occupations, and selected social risks. Simpler models boast high precision whereas recent neural networks improve sensitivity in capturing key entities from free-text fields. Importantly, these NLP approaches recognize more patient social factors than structured EHR data alone to enable richer risk assessments and interventions.

In tandem with advanced informatics solutions, a range of interventions have been tested to address identified social needs and related disparities. These encompass social programs like community health initiatives, resource referrals and patient navigation services, integrated care management models, and group education sessions with peer support. Certain studies have allocated supplementary resources such as medical staff or equipment to vulnerable groups. At times, system-level policy changes are required to promote health equity. Such initiatives have been executed across community, clinic, and health system settings – each with their own merits and challenges. While many efforts underline promising impact on knowledge, self-efficacy, and even select health outcomes, more evidence surrounding sustainability, scalability, and implementation barriers is required.

Moving forward, better standardized corpora are needed to develop and evaluate reusable NLP systems for social domains. Similarly, integration of validated SDoH screening workflows rather than one-off analyses will facilitate routine practice. Ontologies, shareable custom systems, and improved linkages to longitudinal outcomes can further the field. In tandem, more rigorous assessment of multi-sector SDoH interventions and targeting of specific mechanisms of impact will maximize reach across at-risk populations.

### SDoH and Health Outcomes

This review provides useful insights into current approaches and gaps in research on SDoH and health outcomes. Most studies were retrospective analyses examining links between neighborhood disadvantage, food and housing issues, healthcare access barriers and related problems. They studied connections to healthcare use, chronic illness control, and infection diseases. Some assessed mental health, cancer, mortality but less so. Neighborhood factors were studied most among social elements, followed by individual/family-level food and housing problems.

COVID-19 has stimulated greater attention to health disparities and social determinants. Nearly 20 studies examined links between deprivation, barriers, instability and infection rates, severity, and outcomes. Consistently higher COVID-19 risks and deaths among minorities, low-income groups, and those with prior conditions underscored existing inequitable distribution of social risks. Researchers leveraged diverse data from records, census indices, and surveys to quantify the disproportionate pandemic burden on disadvantaged groups. Studies also displayed sophisticated applications of predictive analytics and machine learning to model dynamics. This crisis has expanded SDoH data infrastructure and methodology while underscoring long-term disparities. Assessing pandemic response and recovery across social levels is critical, as disruptions threaten to worsen effects on vulnerable groups.

Techniques like regression modeling helped characterize adjusted outcome associations. But prediction and more advanced analytics were less common. There is some consistent evidence tying greater social deprivation to poorer health across conditions. But differences across settings and mixed findings remain. More controlled, computational research could better uncover precise interactions. Clearer reporting on predictive models applied could also help comparisons to guide health equity solutions.

We see more screening for patient social needs in clinics now. But we still need standardized processes and data integration into records for tracking over time. Currently observational analyses dominate. Moving forward, translating findings into community initiatives for at-risk groups remains vital.

Analyzing Real-World Data in healthcare, especially with the integration of SDoH, is challenging due to the lack of standardized frameworks and complex nature of SDoH data. Developing analytic guidelines is crucial for effectively navigating these complexities, enabling the extraction of actionable insights that inform healthcare decisions and strategies. Such advancements are essential for health outcomes research, as they provide a foundation for creating more effective, evidence-based interventions and policies that consider the broad influences of social factors on health.

### SDoH Interventions

An intervention in response to health care-related issues may transpire at the micro, meso, and macro levels, namely patient care, healthcare institutions, and healthcare policy [243–245]. According to the study, SDoH recognition facilitates intervention programs more at the meso level, such as primary care and specialist referrals, patient navigation services, integrated care management, group education sessions, and medical staff or equipment allocation. A few policy modifications piloted macro-level practice transformations. Only about half of the SDoH-navigated programs emphasized family/peer support and social connections for individual patients (micro-level) as part of the intervention.

The majority of interventions had some benefit, but because they only addressed a few targets in one health system, their generalizability was restricted. This finding holds true in the US, where health delivery systems are fragmented as regional networks. It also demonstrates the challenge of implementing interventions for vulnerable populations [246,247], whose demographics vary across communities on account of cultural and geographical factors. It is imperative for healthcare professionals to bear in mind that identifying SDoH within a community is the initial stage. Establishing connections between individuals who are grappling with health issues and social issues can be challenging due to mental or physical difficulties. The process of establishing trust with vulnerable individuals is iterative and requires ongoing emotional and material input from healthcare professionals in collaboration with community social workers. Besides, in the formation of a regional support network, stakeholders forge lasting partnerships with organizations holding the necessary resources to address challenges identified by SDoH (e.g., accommodation, and transportation).

For more effective intervention, a mature plan with SDoH collection tools embedded in an EHR system should be adopted and modified to reflect the composition and needs of the population being surveyed. The panel should select the platform for survey distribution and storage in order to prevent duplication of effort and data loss and ensure the program’s sustainability. They should ensure that the data utilized to inform interventions is collected directly from the population undergoing the intervention. Utilizing a pilot survey to pre-test a data collection instrument in line with a co-design methodology is suggested [248,249]. Co-designing is a collaborative effort aimed at establishing a conduit for vulnerable individuals to develop a more supportive environment, thereby mitigating the occurrence of unanticipated obstacles.

### Cross-Cutting Insights

Despite some progress, barriers like screening variability [250], data silos [251], predictive model opacity [252], and program adoption challenges [246] restrict reliable, equitable SDoH data usage. Presently, expertise resides in siloes—screening, linkage, extraction, analyses, programs. But real optimization depends on an integrated “full-stack” approach recognizing interdependencies. For example, incorporating extensive SDoH variables strains statistical models in outcomes research, demanding guidance on principled variable selection and penalization procedures. Breaking down walls between activities could promote a comprehensive platform spanning the spectrum from collection to application.

This demands rethinking workflows and breaking down antiquated interdependency barriers that isolate screening from risk detection or hinder integrating contextual data into real-world utilities [26]. Architectural paradigms promoting modular reuse with transparency safeguards can help reconfigure fragmented efforts into an interoperable ecosystem better serving those with disproportionate risk. [253] Since conducting searches, exponential growth in large language models (LLMs) has occurred, presenting new opportunities [254,255]. LLM-based extraction and temporal reasoning models could facilitate reliable SDoH entity recognition across contexts [254]. As research continues, embracing interoperable design principles and controlled evaluation around representative datasets, model transparency, and equitable outcomes remains vital [252].

### Limitations

This scoping review faces certain limitations in comprehensively capturing the state of SDoH data integration into EHRs. Firstly, while efficient, relying solely on PubMed for literature searches and English papers only may introduce selection bias, omitting potentially relevant research indexed in other databases. Supplementing with sources like SCOPUS or Web of Science could have revealed additional insights and applications.

Second, due to resource constraints, the metadata extraction from the final set of included studies was completed by a single reviewer. Having dual independent extraction with consensus meetings is ideal to ensure accuracy and completeness of scoping review data abstractions. The feasibility and impact of implementing a second reader should be evaluated in future updates to strengthen robustness.

Finally, heterogeneity across settings, populations, tools, and outcomes creates complexity in evaluating SDoH-EHR integration maturity. Varying implementation stages and study designs introduce difficulty benchmarking best practices. The scoping methodology prioritized inclusiveness over appraising integration quality, leaving gaps in assessing real-world effectiveness. Capturing nuanced, multi-dimensional integration processes by diverse healthcare systems persists as a challenge, though framework refinement helps structure insights.

## Conclusion

Overall, while collecting patient social contexts shows immense potential to rectify health disparities, realizing possibilities requires ongoing informatics innovation alongside economic investments and policy reforms targeting root societal drivers. This review contributes an evidence base for such continued progress in wisely applying multidimensional SDoH data to promote health equity.

The integration of SDoH data into healthcare practice holds transformative potential for addressing health disparities. Realizing this potential demands continued innovation, strategic investment, and policy evolution, guided by the evidence and insights garnered from comprehensive SDoH research.

## Data Availability

Extracted metadata for all articles is available upon reasonable request to the authors. We will release the data online if published by journal.

## Acknowledgements

We thank the National Institutes of Health for supporting this research: R01-HL162354, K24-HL167127-01A1, and P50MH127511.

## Contributorship Statement

M.J.B. C.L., and D.L.M: Conceptualized the study design and methodology.

C.L., D.L.M., X.M., S.H, and M.J.B.: Led data synthesis and analysis as well as initial manuscript drafting.

C.L., R.Y., S.H., D.L.M., U.V., H.K.D., and P.F.: Contributed to metadata extraction from selected articles.

C.L., Z.A. and H.K.D.: Developed visualizations and tables to summarize key data.

All authors contributed intellectually to the interpretation of findings, critically revised the manuscript, gave final approval of the version to be published, and agree to be accountable for the work.

## Supplement

**Supplement Table 1.**
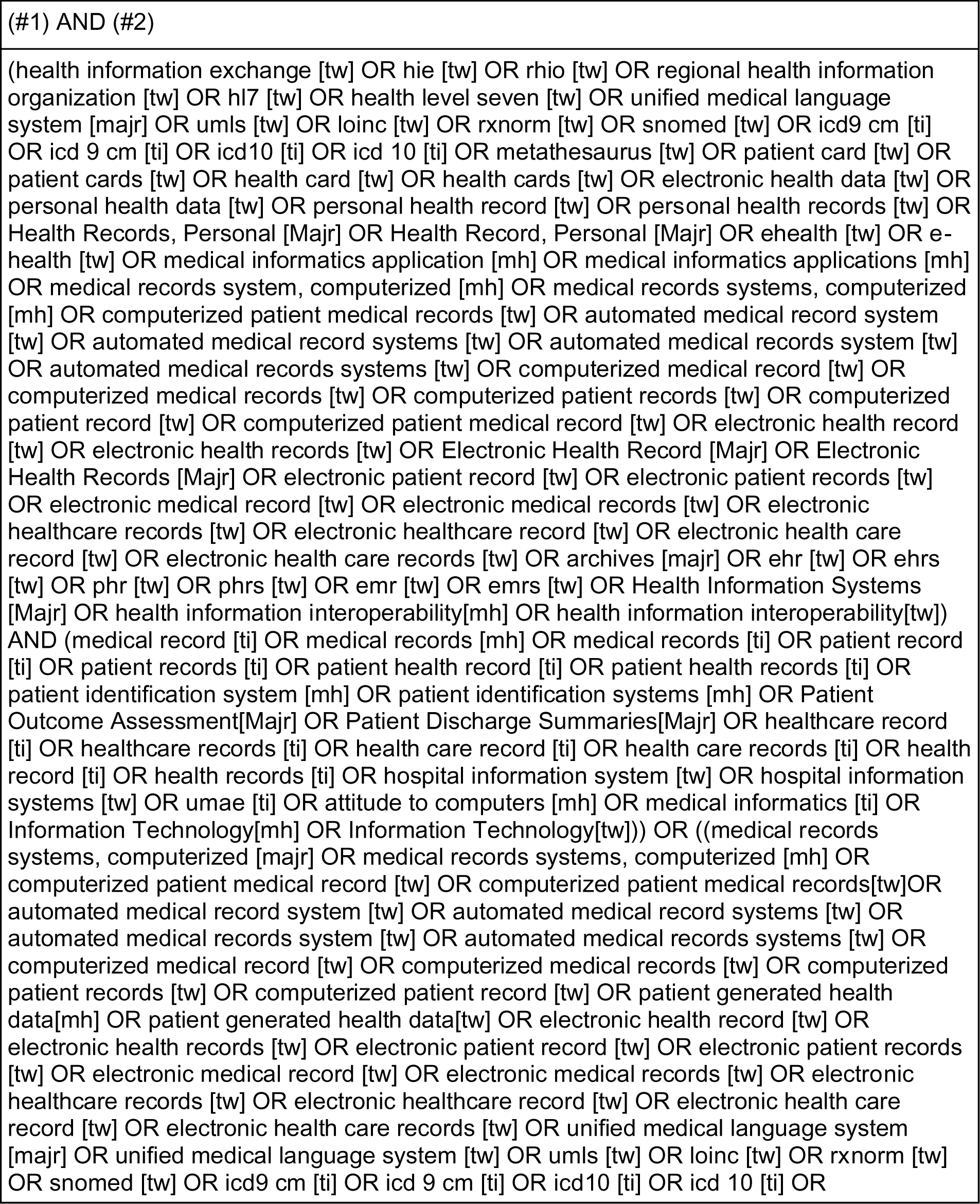
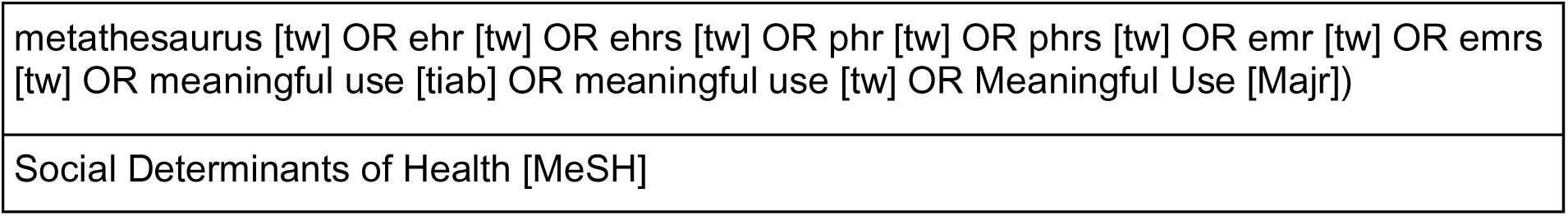
– Search Strategy (2023. May 8th on PubMed)

**Supplement Table 2.** – Screening Included Paper by Domain:

| Domain | Title/Abstract screening |  | Full-text screening |  |
| --- | --- | --- | --- | --- |
|  | Count (sum = 415) | Percentage | (n = 324) | Percentage |
| SDoH and health outcomes | 168 | 40.5% | 164 | 50.6% |
| SDoH data collection and documentation | 132 | 31.8% | 76 | 23.5% |
| NLP for SDoH | 42 | 10.1% | 36 | 11.1% |
| Review Paper | 28 | 6.7% | / | / |
| SDoH screening tools and assessments | 26 | 6.3% | 29 | 9.0% |
| SDoH Intervention | 19 | 4.6% | 19 | 5.9% |

Supplement Table 3 – Meta Data – SDoH Screening and Assessment

Supplement Table 4 – Meta Data – SDoH Data Collection and Documentation

Supplement Table 5 – Meta Data – NLP for SDoH

Supplement Table 6 – Meta Data – SDoH and Health Outcomes

Supplement Table 7 – Meta Data – SDoH Intervention

## References

1. Braveman P. The Social Determinants of Health and Health Disparities. Oxford University Press, 2023.

2. Marmot M. Social determinants of health inequalities. The Lancet 2005; 365(9464): 1099–1104.

3. WHO Commission on Social Determinants of Health, World Health Organization. Closing the Gap in a Generation: Health Equity Through Action on the Social Determinants of Health: Commission on Social Determinants of Health Final Report. World Health Organization, 2008.

4. Social determinants of health. [cited January 26, 2024]. (https://health.gov/healthypeople/priority-areas/social-determinants-health).

5. Barragan, [d-Ca-44] ND. Improving Social Determinants of Health Act of 2021. 379 Feb 2, 2021.

6. Whitman A, De Lew N, Chappel A, Aysola V, Zuckerman R, Sommers BD. Addressing social determinants of health: Examples of successful evidence-based strategies and current federal efforts. [cited January 26, 2024]. (https://www.aspe.hhs.gov/sites/default/files/documents/e2b650cd64cf84aae8ff0fae7474af82/SDOH-Evidence-Review.pdf).

7. Hood CM, Gennuso KP, Swain GR, Catlin BB. County Health Rankings: Relationships Between Determinant Factors and Health Outcomes. American journal of preventive medicine 2016; 50(2): 129–135.

8. Department of Health and Human Services US. physical activity guidelines for Americans (ODPHP Publication No. U0036). Washington, DC: US Government.

9. Hacker K, Auerbach J, Ikeda R, Philip C, Houry D. Social Determinants of Health-An Approach Taken at CDC. Journal of public health management and practice: JPHMP 2022; 28(6): 589–594.

10. PRAPARE. [cited January 31, 2024]. (https://prapare.org/).

11. Rogers CK, Parulekar M, Malik F, Torres CA. A Local Perspective into Electronic Health Record Design, Integration, and Implementation of Screening and Referral for Social Determinants of Health. Perspectives in health information management / AHIMA, American Health Information Management Association 2022; 19(Spring): 1g.

12. Zulman DM, Maciejewski ML, Grubber JM, et al. Patient-Reported Social and Behavioral Determinants of Health and Estimated Risk of Hospitalization in High-Risk Veterans Affairs Patients. JAMA Netw Open 2020; 3(10): e2021457.

13. Dang Y, Li F, Hu X, et al. Systematic design and data-driven evaluation of social determinants of health ontology (SDoHO). Journal of the American Medical Informatics Association: JAMIA 2023; 30(9): 1465–1473.

14. Phuong J, Zampino E, Dobbins N, et al. Extracting Patient-level Social Determinants of Health into the OMOP Common Data Model. AMIA … Annual Symposium proceedings / AMIA Symposium. AMIA Symposium 2021; 2021: 989–998.

15. HL7.FHIR.US.SDOH-CLINICALCARE\gravity value sets – FHIR v4.0.1. [cited January 31, 2024]. (https://build.fhir.org/ig/HL7/fhir-sdoh-clinicalcare/gravity_terminology.html).

16. Kessler RC, Bauer MS, Bishop TM, et al. Evaluation of a Model to Target High-risk Psychiatric Inpatients for an Intensive Postdischarge Suicide Prevention Intervention. JAMA psychiatry 2023; 80(3): 230–240.

17. Gattu RK, Paik G, Wang Y, Ray P, Lichenstein R, Black MM. The Hunger Vital Sign Identifies Household Food Insecurity among Children in Emergency Departments and Primary Care. Children 2019; 6(10) Published online: October 2, 2019. doi:10.3390/children6100107.

18. Johnson D, Saavedra P, Sun E, et al. Community health workers and medicaid managed care in New Mexico. Journal of community health 2012; 37(3): 563–571.

19. Navathe AS, Zhong F, Lei VJ, et al. Hospital Readmission and Social Risk Factors Identified from Physician Notes. Health services research 2018; 53(2): 1110–1136.

20. Amarashingham R, Xie B, Karam A, Nguyen N, Kapoor B. Using Community Partnerships to Integrate Health and Social Services for High-Need, High-Cost Patients. Issue brief 2018; 2018: 1–11.

21. Parmar P, Ryu J, Pandya S, Sedoc J, Agarwal S. Health-focused conversational agents in person-centered care: a review of apps. NPJ digital medicine 2022; 5(1): 21.

22. Gottlieb L, Tobey R, Cantor J, Hessler D, Adler NE. Integrating Social And Medical Data To Improve Population Health: Opportunities And Barriers. Health affairs 2016; 35(11): 2116–2123.

23. Harle CA, Wu W, Vest JR. Accuracy of Electronic Health Record Food Insecurity, Housing Instability, and Financial Strain Screening in Adult Primary Care. JAMA: the journal of the American Medical Association 2023; 329(5): 423–424.

24. Wetta RE, Severin RD, Gruhler H. An evidence-based strategy to achieve equivalency and interoperability for social-behavioral determinants of health assessment, storage, exchange, and use. Health informatics journal 2020; 26(2): 1477–1488.

25. Phuong J, Hong S, Palchuk MB, et al. Advancing Interoperability of Patient-level Social Determinants of Health Data to Support COVID-19 Research. AMIA Joint Summits on Translational Science proceedings. AMIA Joint Summits on Translational Science 2022; 2022: 396–405.

26. Gruß I, Bunce A, Davis J, Dambrun K, Cottrell E, Gold R. Initiating and Implementing Social Determinants of Health Data Collection in Community Health Centers. Population health management 2021; 24(1): 52–58.

27. Council DP. November. The US Playbook to Address Social Determinants Of Health. and Technology Policy. The White House. Retrieved ….

28. Wang M, Pantell MS, Gottlieb LM, Adler-Milstein J. Documentation and review of social determinants of health data in the EHR: measures and associated insights. Journal of the American Medical Informatics Association: JAMIA 2021; 28(12): 2608–2616.

29. Gold R, Bunce A, Cowburn S, et al. Adoption of Social Determinants of Health EHR Tools by Community Health Centers. Annals of family medicine 2018; 16(5): 399–407.

30. Hwang S, Urbanowicz R, Lynch S, et al. Toward Predicting 30-Day Readmission Among Oncology Patients: Identifying Timely and Actionable Risk Factors. JCO clinical cancer informatics 2023; 7: e2200097.

31. Tricco AC, Lillie E, Zarin W, et al. PRISMA Extension for Scoping Reviews (PRISMA-ScR): Checklist and Explanation. Annals of internal medicine 2018; 169(7): 467–473.

32. Graif C, Meurer J, Fontana M. An Ecological Model to Frame the Delivery of Pediatric Preventive Care. Pediatrics 2021; 148(Suppl 1): s13–s20.

33. Bechtel N, Jones A, Kue J, Ford JL. Evaluation of the core 5 social determinants of health screening tool. Public health nursing 2022; 39(2): 438–445.

34. Barton LR, Parke KA, White CL. Screening for the Social and Behavioral Determinants of Health at a School-Based Clinic. Journal of pediatric health care: official publication of National Association of Pediatric Nurse Associates & Practitioners 2019; 33(5): 537–544.

35. Gore E, DiTursi J, Rambuss R, Pope-Collins E, Train MK. Implementing a Process for Screening Hospitalized Adults for Food Insecurity at a Tertiary Care Center. Journal for healthcare quality: official publication of the National Association for Healthcare Quality 2022; 44(5): 305–312.

36. Vasan A, Kenyon CC, Palakshappa D. Differences in Pediatric Residents’ Social Needs Screening Practices Across Health Care Settings. Hospital pediatrics 2020; 10(5): 443– 446.

37. Berkowitz RL, Bui L, Shen Z, et al. Evaluation of a social determinants of health screening questionnaire and workflow pilot within an adult ambulatory clinic. BMC family practice 2021; 22(1): 256.

38. Meyer D, Lerner E, Phillips A, Zumwalt K. Universal Screening of Social Determinants of Health at a Large US Academic Medical Center, 2018. American journal of public health 2020; 110(S2): S219–S221.

39. Nohria R, Xiao N, Guardado R, et al. Implementing Health Related Social Needs Screening in an Outpatient Clinic. Journal of primary care & community health 2022; 13: 21501319221118809.

40. Wallace AS, Luther B, Guo J-W, Wang C-Y, Sisler S, Wong B. Implementing a Social Determinants Screening and Referral Infrastructure During Routine Emergency Department Visits, Utah, 2017-2018. Preventing chronic disease 2020; 17: E45.

41. Hatef E, Rouhizadeh M, Tia I, et al. Assessing the Availability of Data on Social and Behavioral Determinants in Structured and Unstructured Electronic Health Records: A Retrospective Analysis of a Multilevel Health Care System. JMIR medical informatics 2019; 7(3): e13802.

42. Chukmaitov A, Dahman B, Garland SL, et al. Addressing social risk factors in the inpatient setting: Initial findings from a screening and referral pilot at an urban safety-net academic medical center in Virginia, USA. Preventive medicine reports 2022; 29: 101935.

43. Yazar V, Kang S-U, Ha S, Dawson VL, Dawson TM. Integrative genome-wide analysis of dopaminergic neuron-specific PARIS expression in Drosophila dissects recognition of multiple PPAR-γ associated gene regulation. Scientific reports 2021; 11(1): 21500.

44. Okafor M, Chiu S, Feinn R. Quantitative and qualitative results from implementation of a two-item food insecurity screening tool in healthcare settings in Connecticut. Preventive medicine reports 2020; 20: 101191.

45. Herrera T, Fiori KP, Archer-Dyer H, Lounsbury DW, Wylie-Rosett J. Social Determinants of Health Screening by Preclinical Medical Students During the COVID-19 Pandemic: Service-Based Learning Case Study. JMIR medical education 2022; 8(1): e32818.

46. Potharaju KA, Fields JD, Cemballi AG, et al. Assessing Alignment of Patient and Clinician Perspectives on Community Health Resources for Chronic Disease Management. Healthcare (Basel, Switzerland) 2022; 10(10)Published online: October 12, 2022. doi:10.3390/healthcare10102006.

47. Vale MD, Perkins DW. Discuss and remember: Clinician strategies for integrating social determinants of health in patient records and care. Social science & medicine 2022; 315: 115548.

48. Senteio C, Adler-Milstein J, Richardson C, Veinot0 T. Psychosocial information use for clinical decisions in diabetes care. Journal of the American Medical Informatics Association: JAMIA 2019; 26(8-9): 813–824.

49. Hirsch A, Durden TE, Silva J. Linking Electronic Health Records and In-Depth Interviews to Inform Efforts to Integrate Social Determinants of Health Into Health Care Delivery: Protocol for a Qualitative Research Study. JMIR research protocols 2022; 11(3): e36201.

50. Lasser EC, Kim JM, Hatef E, Kharrazi H, Marsteller JA, DeCamp LR. Social and Behavioral Variables in the Electronic Health Record: A Path Forward to Increase Data Quality and Utility. Academic medicine: journal of the Association of American Medical Colleges 2021; 96(7): 1050–1056.

51. Vatani H, Sharma H, Azhar K, Kochendorfer KM, Valenta AL, Dunn Lopez K. Required data elements for interprofessional rounds through the lens of multiple professions. Journal of interprofessional care 2020;: 1–7.

52. Nguyen OK, Chan CV, Makam A, Stieglitz H, Amarasingham R. Envisioning a social-health information exchange as a platform to support a patient-centered medical neighborhood: a feasibility study. Journal of general internal medicine 2015; 30(1): 60–67.

53. Lindau ST, Makelarski J, Abramsohn E, et al. CommunityRx: A Population Health Improvement Innovation That Connects Clinics To Communities. Health affairs 2016; 35(11): 2020–2029.

54. Duberstein ZT, Brunner J, Panisch LS, et al. The Biopsychosocial Model and Perinatal Health Care: Determinants of Perinatal Care in a Community Sample. Frontiers in psychiatry / Frontiers Research Foundation 2021; 12: 746803.

55. Duncan PW, Abbott RM, Rushing S, et al. COMPASS-CP: An Electronic Application to Capture Patient-Reported Outcomes to Develop Actionable Stroke and Transient Ischemic Attack Care Plans. Circulation. Cardiovascular quality and outcomes 2018; 11(8): e004444.

56. Gold R, Bunce A, Cottrell E, et al. Study protocol: a pragmatic, stepped-wedge trial of tailored support for implementing social determinants of health documentation/action in community health centers, with realist evaluation. Implementation science: IS 2019; 14(1): 9.

57. Palakshappa D, Benefield AJ, Furgurson KF, et al. Feasibility of Mobile Technology to Identify and Address Patients’ Unmet Social Needs in a Primary Care Clinic. Population health management 2021; 24(3): 385–392.

58. Sposito RS, Selleck C. Expanding Data Reporting Capacity of Free and Charitable Clinics: A Quality Improvement Project. Journal of doctoral nursing practice 2020; 13(1): 64–70.

59. Sitapati AM, Berkovich B, Arellano AM, et al. A case study of the 1115 waiver using population health informatics to address disparities. JAMIA open 2020; 3(2): 178–184.

60. Van Brunt D. Community Health Records: Establishing a Systematic Approach to Improving Social and Physical Determinants of Health. American journal of public health 2017; 107(3): 407–412.

61. Arevian AC, Springgate B, Jones F, et al. The Community and Patient Partnered Research Network (CPPRN): Application of Patient-Centered Outcomes Research to Promote Behavioral Health Equity. Ethnicity & disease 2018; 28(Suppl 2): 295–302.

62. Melton GB, Manaktala S, Sarkar IN, Chen ES. Social and behavioral history information in public health datasets. AMIA … Annual Symposium proceedings / AMIA Symposium. AMIA Symposium 2012; 2012: 625–634.

63. Aghdaee M, Gu Y, Sinha K, Parkinson B, Sharma R, Cutler H. Mapping the Patient-Reported Outcomes Measurement Information System (PROMIS-29) to EQ-5D-5L. PharmacoEconomics 2023; 41(2): 187–198.

64. National Survey of Children’s Health – Data Resource Center for Child and Adolescent Health. [cited February 1, 2024]. (https://www.childhealthdata.org/learn-about-the-nsch/NSCH).

65. Ettinger AK, Landsittel D, Abebe KZ, et al. THRIVE Conceptual Framework and Study Protocol: A Community-Partnered Longitudinal Multi-Cohort Study to Promote Child and Youth Thriving, Health Equity, and Community Strength. Frontiers in pediatrics 2021; 9: 797526.

66. Trinacty CM, LaWall E, Ashton M, Taira D, Seto TB, Sentell T. Adding Social Determinants in the Electronic Health Record in Clinical Care in Hawai’i: Supporting Community-Clinical Linkages in Patient Care. Hawai’i journal of medicine & public health: a journal of Asia Pacific Medicine & Public Health 2019; 78(6 Suppl 1): 46–51.

67. Cottrell EK, Hendricks M, Dambrun K, et al. Comparison of Community-Level and Patient-Level Social Risk Data in a Network of Community Health Centers. JAMA network open 2020; 3(10): e2016852.

68. Gardner BJ, Pedersen JG, Campbell ME, McClay JC. Incorporating a location-based socioeconomic index into a de-identified i2b2 clinical data warehouse. Journal of the American Medical Informatics Association: JAMIA 2019; 26(4): 286–293.

69. Korzeniewski SJ, Bezold C, Carbone JT, et al. The Population Health OutcomEs aNd Information EXchange (PHOENIX) Program – A Transformative Approach to Reduce the Burden of Chronic Disease. Online journal of public health informatics 2020; 12(1): e3.

70. Udalova V, Carey TS, Chelminski PR, et al. Linking Electronic Health Records to the American Community Survey: Feasibility and Process. American journal of public health 2022; 112(6): 923–930.

71. Sacco SJ, Chen K, Wang F, Aseltine R. Target-based fusion using social determinants of health to enhance suicide prediction with electronic health records. PloS one 2023; 18(4): e0283595.

72. Gottlieb LM, Tirozzi KJ, Manchanda R, Burns AR, Sandel MT. Moving electronic medical records upstream: incorporating social determinants of health. American journal of preventive medicine 2015; 48(2): 215–218.

73. McCormack LA, Madlock-Brown C. Social Determinant of Health Documentation Trends and Their Association with Emergency Department Admissions. AMIA … Annual Symposium proceedings / AMIA Symposium. AMIA Symposium 2020; 2020: 823–832.

74. Hewner S, Casucci S, Sullivan S, et al. Integrating Social Determinants of Health into Primary Care Clinical and Informational Workflow during Care Transitions. EGEMS (Washington, DC) 2017; 5(2): 2.

75. Dusetzina SBPhD, Enewold LMph PhD, Gentile DPhD, Ramsey SDMd PhD, Halpern MT. New Data Resources, Linkages, and Infrastructure for Cancer Health Economics Research: Main Topics From a Panel Discussion. Journal of the National Cancer Institute. Monographs 2022; 2022(59): 68–73.

76. Kane NJ, Wang X, Gerkovich MM, et al. The Envirome Web Service: Patient context at the point of care. Journal of biomedical informatics 2021; 119: 103817.

77. Biro S, Williamson T, Leggett JA, et al. Utility of linking primary care electronic medical records with Canadian census data to study the determinants of chronic disease: an example based on socioeconomic status and obesity. BMC medical informatics and decision making 2016; 16: 32.

78. Comer KF, Grannis S, Dixon BE, Bodenhamer DJ, Wiehe SE. Incorporating geospatial capacity within clinical data systems to address social determinants of health. Public health reports 2011; 126 Suppl 3(Suppl 3): 54–61.

79. Bambekova PG, Liaw W, Phillips RL Jr, Bazemore A. Integrating Community and Clinical Data to Assess Patient Risks with A Population Health Assessment Engine (PHATE). Journal of the American Board of Family Medicine: JABFM 2020; 33(3): 463– 467.

80. Hughes LS, Phillips RL Jr, DeVoe JE, Bazemore AW. Community Vital Signs: Taking the Pulse of the Community While Caring for Patients. Journal of the American Board of Family Medicine: JABFM 2016; 29(3): 419–422.

81. Bazemore AW, Cottrell EK, Gold R, et al. “Community vital signs”: incorporating geocoded social determinants into electronic records to promote patient and population health. Journal of the American Medical Informatics Association: JAMIA 2016; 23(2): 407– 412.

82. Buitron de la Vega P, Losi S, Sprague Martinez L, et al. Implementing an EHR-based Screening and Referral System to Address Social Determinants of Health in Primary Care. Medical care 2019; 57 Suppl 6 Suppl 2: S133–S139.

83. Luijks H, van Boven K, Olde Hartman T, Uijen A, van Weel C, Schers H. Purposeful Incorporation of Patient Narratives in the Medical Record in the Netherlands. Journal of the American Board of Family Medicine: JABFM 2021; 34(4): 709–723.

84. Lee TC, Saseendrakumar BR, Nayak M, et al. Social Determinants of Health Data Availability for Patients with Eye Conditions. Ophthalmology science 2022; 2(2) Published online: June 2022. doi:10.1016/j.xops.2022.100151.

85. Jain A, van Hoek AJ, Walker JL, Mathur R, Smeeth L, Thomas SL. Identifying social factors amongst older individuals in linked electronic health records: An assessment in a population based study. PloS one 2017; 12(11): e0189038.

86. Rousseau JF, Oliveira E, Tierney WM, Khurshid A. Methods for development and application of data standards in an ontology-driven information model for measuring, managing, and computing social determinants of health for individuals, households, and communities evaluated through an example of asthma. Journal of biomedical informatics 2022; 136: 104241.

87. Stewart de Ramirez S, Shallat J, McClure K, Foulger R, Barenblat L. Screening for Social Determinants of Health: Active and Passive Information Retrieval Methods. Population health management 2022; 25(6): 781–788.

88. Tan-McGrory A, Bennett-AbuAyyash C, Gee S, et al. A patient and family data domain collection framework for identifying disparities in pediatrics: results from the pediatric health equity collaborative. BMC pediatrics 2018; 18(1): 18.

89. Kepper MM, Walsh-Bailey C, Prusaczyk B, Zhao M, Herrick C, Foraker R. The adoption of social determinants of health documentation in clinical settings. Health services research 2023; 58(1): 67–77.

90. Yang X, Yelton B, Chen S, et al. Examining Social Determinants of Health During a Pandemic: Clinical Application of Z Codes Before and During COVID-19. Frontiers in public health 2022; 10: 888459.

91. Richman EL, Lombardi BM, de Saxe Zerden L, Forte AB. What Do EHRs Tell Us about How We Deploy Health Professionals to Address the Social Determinants of Health. Social work in public health 2022; 37(3): 287–296.

92. Albert SM, McCracken P, Bui T, et al. Do patients want clinicians to ask about social needs and include this information in their medical record? BMC health services research 2022; 22(1): 1275.

93. Freij M, Dullabh P, Lewis S, Smith SR, Hovey L, Dhopeshwarkar R. Incorporating Social Determinants of Health in Electronic Health Records: Qualitative Study of Current Practices Among Top Vendors. JMIR medical informatics 2019; 7(2): e13849.

94. Tenenbaum JD, Christian V, Cornish MA, et al. The MURDOCK Study: a long-term initiative for disease reclassification through advanced biomarker discovery and integration with electronic health records. American journal of translational research 2012; 4(3): 291– 301.

95. Shi Q, Herbert C, Ward DV, et al. COVID-19 Variant Surveillance and Social Determinants in Central Massachusetts: Development Study. JMIR formative research 2022; 6(6): e37858.

96. Ofili EO, Schanberg LE, Hutchinson B, et al. The Association of Black Cardiologists (ABC) Cardiovascular Implementation Study (CVIS): A Research Registry Integrating Social Determinants to Support Care for Underserved Patients. International journal of environmental research and public health 2019; 16(9) Published online: May 10, 2019. doi:10.3390/ijerph16091631.

97. Kyriazis D, Autexier S, Brondino I, et al. CrowdHEALTH: Holistic Health Records and Big Data Analytics for Health Policy Making and Personalized Health. Studies in health technology and informatics 2017; 238: 19–23.

98. Khatib R, Li Y, Glowacki N, Siddiqi A. Addressing Social Needs in the Clinical Setting: Description of Needs Identified in a Quality Improvement Pilot Across 3 Community Hospital Service Areas. Population health management 2022; 25(5): 632–638.

99. Smith MA, Gigot M, Harburn A, et al. Insights into measuring health disparities using electronic health records from a statewide network of health systems: A case study. Journal of clinical and translational science 2023; 7(1): e54.

100. Cook L, Espinoza J, Weiskopf NG, et al. Issues With Variability in Electronic Health Record Data About Race and Ethnicity: Descriptive Analysis of the National COVID Cohort Collaborative Data Enclave. JMIR medical informatics 2022; 10(9): e39235.

101. Winden TJ, Chen ES, Melton GB. Representing Residence, Living Situation, and Living Conditions: An Evaluation of Terminologies, Standards, Guidelines, and Measures/Surveys. AMIA … Annual Symposium proceedings / AMIA Symposium. AMIA Symposium 2016; 2016: 2072–2081.

102. Lindemann EA, Chen ES, Wang Y, Skube SJ, Melton GB. Representation of Social History Factors Across Age Groups: A Topic Analysis of Free-Text Social Documentation. AMIA … Annual Symposium proceedings / AMIA Symposium. AMIA Symposium 2017; 2017: 1169–1178.

103. Dorr DA, Quiñones AR, King T, Wei MY, White K, Bejan CA. Prediction of Future Health Care Utilization Through Note-extracted Psychosocial Factors. Medical care 2022; 60(8): 570–578.

104. Bejan CA, Angiolillo J, Conway D, et al. Mining 100 million notes to find homelessness and adverse childhood experiences: 2 case studies of rare and severe social determinants of health in electronic health records. Journal of the American Medical Informatics Association: JAMIA 2018; 25(1): 61–71.

105. Dorr D, Bejan CA, Pizzimenti C, Singh S, Storer M, Quinones A. Identifying Patients with Significant Problems Related to Social Determinants of Health with Natural Language Processing. Studies in health technology and informatics 2019; 264: 1456–1457.

106. Conway M, Keyhani S, Christensen L, et al. Moonstone: a novel natural language processing system for inferring social risk from clinical narratives. Journal of biomedical semantics 2019; 10(1): 6.

107. Bucher BT, Shi J, Pettit RJ, Ferraro J, Chapman WW, Gundlapalli A. Determination of Marital Status of Patients from Structured and Unstructured Electronic Healthcare Data. AMIA … Annual Symposium proceedings / AMIA Symposium. AMIA Symposium 2019; 2019: 267–274.

108. Oreskovic NM, Maniates J, Weilburg J, Choy G. Optimizing the Use of Electronic Health Records to Identify High-Risk Psychosocial Determinants of Health. JMIR medical informatics 2017; 5(3): e25.

109. Morrow D, Zamora-Resendiz R, Beckham JC, et al. A case for developing domain-specific vocabularies for extracting suicide factors from healthcare notes. Journal of psychiatric research 2022; 151: 328–338.

110. Chilman N, Song X, Roberts A, et al. Text mining occupations from the mental health electronic health record: a natural language processing approach using records from the Clinical Record Interactive Search (CRIS) platform in south London, UK. BMJ open 2021; 11(3): e042274.

111. Rawat BPS, Keating H, Goodwin R, Druhl E, Yu H. An Investigation of the Representation of Social Determinants of Health in the UMLS. AMIA … Annual Symposium proceedings / AMIA Symposium. AMIA Symposium 2022; 2022: 912–921.

112. Shah-Mohammadi F, Cui W, Bachi K, Hurd Y, Finkelstein J. Comparative Analysis of Patient Distress in Opioid Treatment Programs using Natural Language Processing. Biomedical engineering systems and technologies, international joint conference, BIOSTEC … revised selected papers. BIOSTEC (Conference) 2022; 2022: 319–326.

113. Wang EA, Long JB, McGinnis KA, et al. Measuring Exposure to Incarceration Using the Electronic Health Record. Medical care 2019; 57 Suppl 6 Suppl 2(Suppl 6 2): S157–S163.

114. Hatef E, Rouhizadeh M, Nau C, et al. Development and assessment of a natural language processing model to identify residential instability in electronic health records’ unstructured data: a comparison of 3 integrated healthcare delivery systems. JAMIA open 2022; 5(1): ooac006.

115. Hatef E, Singh Deol G, Rouhizadeh M, et al. Measuring the Value of a Practical Text Mining Approach to Identify Patients With Housing Issues in the Free-Text Notes in Electronic Health Record: Findings of a Retrospective Cohort Study. Frontiers in public health 2021; 9: 697501.

116. Chapman AB, Jones A, Kelley AT, et al. ReHouSED: A novel measurement of Veteran housing stability using natural language processing. Journal of biomedical informatics 2021; 122: 103903.

117. Feller DJ, Bear Don’t Walk OJ Iv, Zucker J, Yin MT, Gordon P, Elhadad N. Detecting Social and Behavioral Determinants of Health with Structured and Free-Text Clinical Data. Applied clinical informatics 2020; 11(1): 172–181.

118. Ahsan H, Ohnuki E, Mitra A, Yu H. MIMIC-SBDH: A Dataset for Social and Behavioral Determinants of Health. Proceedings of machine learning research 2021; 149: 391–413.

119. Feller DJ, Zucker J, Yin MT, Gordon P, Elhadad N. Using Clinical Notes and Natural Language Processing for Automated HIV Risk Assessment. Journal of acquired immune deficiency syndromes 2018; 77(2): 160–166.

120. Rouillard CJ, Nasser MA, Hu H, Roblin DW. Evaluation of a Natural Language Processing Approach to Identify Social Determinants of Health in Electronic Health Records in a Diverse Community Cohort. Medical care 2022; 60(3): 248–255.

121. Teng A, Wilcox A. Simplified data science approach to extract social and behavioural determinants: a retrospective chart review. BMJ open 2022; 12(1): e048397.

122. Yu Z, Yang X, Guo Y, Bian J, Wu Y. Assessing the Documentation of Social Determinants of Health for Lung Cancer Patients in Clinical Narratives. Frontiers in public health 2022; 10: 778463.

123. Yu Z, Yang X, Dang C, et al. A Study of Social and Behavioral Determinants of Health in Lung Cancer Patients Using Transformers-based Natural Language Processing Models. AMIA … Annual Symposium proceedings / AMIA Symposium. AMIA Symposium 2021; 2021: 1225–1233.

124. Bashir SR, Raza S, Kocaman V, Qamar U. Clinical Application of Detecting COVID-19 Risks: A Natural Language Processing Approach. Viruses 2022; 14(12) Published online: December 11, 2022. doi:10.3390/v14122761.

125. Mitra A, Pradhan R, Melamed RD, et al. Associations Between Natural Language Processing-Enriched Social Determinants of Health and Suicide Death Among US Veterans. JAMA network open 2023; 6(3): e233079.

126. Richie R, Ruiz VM, Han S, Shi L, Tsui FR. Extracting social determinants of health events with transformer-based multitask, multilabel named entity recognition. Journal of the American Medical Informatics Association: JAMIA 2023; 30(8): 1379–1388.

127. Zhao X, Rios A. A marker-based neural network system for extracting social determinants of health. Journal of the American Medical Informatics Association: JAMIA 2023; 30(8): 1398–1407.

128. Newman-Griffis D, Fosler-Lussier E. Automated Coding of Under-Studied Medical Concept Domains: Linking Physical Activity Reports to the International Classification of Functioning, Disability, and Health. Frontiers in digital health 2021; 3 Published online: March 2021. doi:10.3389/fdgth.2021.620828.

129. Mitra A, Ahsan H, Li W, et al. Risk Factors Associated With Nonfatal Opioid Overdose Leading to Intensive Care Unit Admission: A Cross-sectional Study. JMIR medical informatics 2021; 9(11): e32851.

130. Lybarger K, Dobbins NJ, Long R, et al. Leveraging natural language processing to augment structured social determinants of health data in the electronic health record. Journal of the American Medical Informatics Association: JAMIA 2023; 30(8): 1389–1397.

131. Han S, Zhang RF, Shi L, et al. Classifying social determinants of health from unstructured electronic health records using deep learning-based natural language processing. Journal of biomedical informatics 2022; 127: 103984.

132. Wong L, Yu F, Bhattacharyya S, Greer ML. Covid-19 Positivity Differences Among Patients of a Rural, Southern US State Hospital System Based on Population Density, Rural-Urban Classification, and Area Deprivation Index. Studies in health technology and informatics 2022; 294: 701–702.

133. Giovanatti A, Elassar H, Karabon P, Wunderlich-Barillas T, Halalau A. Social Determinants of Health Correlating with Mechanical Ventilation of COVID-19 Patients: A Multi-Center Observational Study. International journal of general medicine 2021; 14: 8521–8526.

134. David P, Fracci S, Wojtowicz J, et al. Ethnicity, Social Determinants of Health, and Pediatric Primary Care During the COVID-19 Pandemic. Journal of primary care & community health 2022; 13: 21501319221112248.

135. Harding JL, Patel SA, Davis T, et al. Understanding Racial Disparities in COVID-19-Related Complications: Protocol for a Mixed Methods Study. JMIR research protocols 2022; 11(10): e38914.

136. Holbert SE, Andersen K, Stone D, Pipkin K, Turcotte J, Patton C. Social Determinants of Health Influence Early Outcomes Following Lumbar Spine Surgery. The Ochsner journal 2022; 22(4): 299–306.

137. Ye Y, Beachy MW, Luo J, et al. Geospatial, Clinical, and Social Determinants of Hospital Readmissions. American journal of medical quality: the official journal of the American College of Medical Quality 2019; 34(6): 607–614.

138. Johnson AE, Zhu J, Garrard W, et al. Area Deprivation Index and Cardiac Readmissions: Evaluating Risk-Prediction in an Electronic Health Record. Journal of the American Heart Association 2021; 10(13): e020466.

139. Hall AG, Davlyatov GK, Orewa GN, Mehta TS, Feldman SS. Multiple Electronic Health Record-Based Measures of Social Determinants of Health to Predict Return to the Emergency Department Following Discharge. Population health management 2022; 25(6): 771–780.

140. Jamei M, Nisnevich A, Wetchler E, Sudat S, Liu E. Predicting all-cause risk of 30-day hospital readmission using artificial neural networks. PloS one 2017; 12(7): e0181173.

141. Zhang Y, Zhang Y, Sholle E, et al. Assessing the impact of social determinants of health on predictive models for potentially avoidable 30-day readmission or death. PloS one 2020; 15(6): e0235064.

142. Nijhawan AE, Metsch LR, Zhang S, et al. Clinical and Sociobehavioral Prediction Model of 30-Day Hospital Readmissions Among People With HIV and Substance Use Disorder: Beyond Electronic Health Record Data. Journal of acquired immune deficiency syndromes 2019; 80(3): 330–341.

143. Ehwerhemuepha L, Pugh K, Grant A, et al. A Statistical-Learning Model for Unplanned 7-Day Readmission in Pediatrics. Hospital pediatrics 2020; 10(1): 43–51.

144. Graham LA, Hawn MT, Dasinger EA, et al. Psychosocial Determinants of Readmission After Surgery. Medical care 2021; 59(10): 864–871.

145. Sills MR, Hall M, Cutler GJ, et al. Adding Social Determinant Data Changes Children’s Hospitals’ Readmissions Performance. The Journal of pediatrics 2017; 186: 150–157.e1.

146. Tomayko EJ, Flood TL, Tandias A, Hanrahan LP. Linking electronic health records with community-level data to understand childhood obesity risk. Pediatric obesity 2015; 10(6): 436–441.

147. Prather AA, Gottlieb LM, Giuse NB, et al. National Academy of Medicine Social and Behavioral Measures: Associations With Self-Reported Health. American journal of preventive medicine 2017; 53(4): 449–456.

148. Winckler B, Nguyen M, Khare M, et al. Geographic Variation in Acute Pediatric Mental Health Utilization. Academic pediatrics 2023; 23(2): 448–456.

149. Tai-Seale M, Cheung MW, Kwak J, et al. Unmet needs for food, medicine, and mental health services among vulnerable older adults during the COVID-19 pandemic. Health services research 2023; 58 Suppl 1(Suppl 1): 69–77.

150. Schlauch KA, Read RW, Koning SM, Neveux I, Grzymski JJ. Using phenome-wide association studies and the SF-12 quality of life metric to identify profound consequences of adverse childhood experiences on adult mental and physical health in a Northern Nevadan population. Frontiers in psychiatry / Frontiers Research Foundation 2022; 13: 984366.

151. Avalos LA, Nance N, Zhu Y, et al. Contributions of COVID-19 Pandemic-Related Stressors to Racial and Ethnic Disparities in Mental Health During Pregnancy. Frontiers in psychiatry / Frontiers Research Foundation 2022; 13: 837659.

152. Ben-Assuli O, Vest JR. Return visits to the emergency department: An analysis using group based curve models. Health informatics journal 2022; 28(2): 14604582221105444.

153. Davis CI, Montgomery AE, Dichter ME, Taylor LD, Blosnich JR. Social determinants and emergency department utilization: Findings from the Veterans Health Administration. The American journal of emergency medicine 2020; 38(9): 1904–1909.

154. Mosen DM, Banegas MP, Benuzillo JG, Hu WR, Brooks NB, Ertz-Berger BL. Association Between Social and Economic Needs With Future Healthcare Utilization. American journal of preventive medicine 2020; 58(3): 457–460.

155. Lawson NR, Klein MD, Ollberding NJ, Wurster Ovalle V, Beck AF. The Impact of Infant Well-Child Care Compliance and Social Risks on Emergency Department Utilization. Clinical pediatrics 2017; 56(10): 920–927.

156. Headen IE, Dubbin L, Canchola AJ, Kersten E, Yen IH. Health care utilization among women of reproductive age living in public housing: Associations across six public housing sites in San Francisco. Preventive medicine reports 2022; 27: 101797.

157. Vest JR, Ben-Assuli O. Prediction of emergency department revisits using area-level social determinants of health measures and health information exchange information. International journal of medical informatics 2019; 129: 205–210.

158. Bhavsar NA, Gao A, Phelan M, Pagidipati NJ, Goldstein BA. Value of Neighborhood Socioeconomic Status in Predicting Risk of Outcomes in Studies That Use Electronic Health Record Data. JAMA network open 2018; 1(5): e182716.

159. Faison K, Moon A, Buckman C, et al. Change of address as a measure of housing insecurity predicting rural emergency department revisits after asthma exacerbation. The Journal of asthma: official journal of the Association for the Care of Asthma 2021; 58(12): 1616–1622.

160. Grinspan ZM, Patel AD, Hafeez B, Abramson EL, Kern LM. Predicting frequent emergency department use among children with epilepsy: A retrospective cohort study using electronic health data from 2 centers. Epilepsia 2018; 59(1): 155–169.

161. Weigert RM, McMichael BS, VanderVelden HA, et al. Parental Childhood Adversity and Pediatric Emergency Department Utilization: A Pilot Study. Pediatric emergency care 2022; 38(12): 665–671.

162. Power-Hays A, Patterson A, Sobota A. Household material hardships impact emergency department reliance in pediatric patients with sickle cell disease. Pediatric blood & cancer 2020; 67(10): e28587.

163. Heinert SW, McCoy J, Strickland PO, Riggs R, Eisenstein R. More Accessible COVID-19 Treatment Through Monoclonal Antibody Infusion in the Emergency Department. The western journal of emergency medicine 2022; 23(5): 618–622.

164. Mayfield CA, de Hernandez BU, Geraci M, Eberth JM, Dulin M, Merchant AT. Residential Segregation and Emergency Department Utilization Among an Underserved Urban Emergency Department Sample in North Carolina. North Carolina medical journal 2022; 83(1): 48–57.

165. O’Malley JA, Klett BM, Klein MD, Inman N, Beck AF. Revealing the Prevalence and Consequences of Food Insecurity in Children with Epilepsy. Journal of community health 2017; 42(6): 1213–1219.

166. Goyal P, Schenck E, Wu Y, et al. Influence of social deprivation index on in-hospital outcomes of COVID-19. Scientific reports 2023; 13(1): 1746.

167. Hong AS, Nguyen DQ, Lee SC, et al. Prior Frequent Emergency Department Use as a Predictor of Emergency Department Visits After a New Cancer Diagnosis. JCO oncology practice 2021; 17(11): e1738–e1752.

168. Conroy K, Samnaliev M, Cheek S, Chien AT. Pediatric Primary Care-Based Social Needs Services and Health Care Utilization. Academic pediatrics 2021; 21(8): 1331–1337.

169. Luo J, Tong L, Crotty BH, et al. Telemedicine Adoption during the COVID-19 Pandemic: Gaps and Inequalities. Applied clinical informatics 2021; 12(4): 836–844.

170. Schwartz BS, Kolak M, Pollak JS, et al. Associations of four indexes of social determinants of health and two community typologies with new onset type 2 diabetes across a diverse geography in Pennsylvania. PloS one 2022; 17(9): e0274758.

171. Scarton L, Nelson T, Yao Y, et al. Medication Adherence and Cardiometabolic Control Indicators Among American Indian Adults Receiving Tribal Health Services: Protocol for a Longitudinal Electronic Health Records Study. JMIR research protocols 2022; 11(10): e39193.

172. Li Y, Hu H, Zheng Y, et al. Impact of Contextual-Level Social Determinants of Health on Newer Antidiabetic Drug Adoption in Patients with Type 2 Diabetes. International journal of environmental research and public health 2023; 20(5) Published online: February 24, 2023. doi:10.3390/ijerph20054036.

173. Lê-Scherban F, Ballester L, Castro JC, et al. Identifying neighborhood characteristics associated with diabetes and hypertension control in an urban African-American population using geo-linked electronic health records. Preventive medicine reports 2019; 15: 100953.

174. Kivimäki M, Vahtera J, Tabák AG, et al. Neighbourhood socioeconomic disadvantage, risk factors, and diabetes from childhood to middle age in the Young Finns Study: a cohort study. The Lancet. Public health 2018; 3(8): e365–e373.

175. Cereijo L, Gullón P, Del Cura I, et al. Exercise facilities and the prevalence of obesity and type 2 diabetes in the city of Madrid. Diabetologia 2022; 65(1): 150–158.

176. Brown AGM, Kressin N, Terrin N, et al. The Influence of Health Insurance Stability on Racial/Ethnic Differences in Diabetes Control and Management. Ethnicity & disease 2021; 31(1): 149–158.

177. Matsumoto C-L, Tobe S, Schreiber YS, et al. Diabetes prevalence and demographics in 25 First Nations communities in northwest Ontario (2014-2017). Canadian journal of rural medicine: the official journal of the Society of Rural Physicians of Canada = Journal canadien de la medecine rurale: le journal officiel de la Societe de medecine rurale du Canada 2020; 25(4): 139–144.

178. Cereijo L, Gullón P, Del Cura I, et al. Exercise facility availability and incidence of type 2 diabetes and complications in Spain: A population-based retrospective cohort 2015-2018. Health & place 2023; 81: 103027.

179. Nieuwenhuijse EA, Struijs JN, Sutch SP, Numans ME, Vos RC. Achieving diabetes treatment targets in people with registered mental illness is similar or improved compared with those without: Analyses of linked observational datasets. Diabetic medicine: a journal of the British Diabetic Association 2022; 39(6): e14835.

180. Ares-Blanco S, Polentinos-Castro E, Rodríguez-Cabrera F, Gullón P, Franco M, Del Cura-González I. Inequalities in glycemic and multifactorial cardiovascular control of type 2 diabetes: The Heart Healthy Hoods study. Frontiers of medicine 2022; 9: 966368.

181. Ye C, Fu T, Hao S, et al. Prediction of Incident Hypertension Within the Next Year: Prospective Study Using Statewide Electronic Health Records and Machine Learning. Journal of medical Internet research 2018; 20(1): e22.

182. Sonnenblick R, Reilly A, Roye K, et al. Social Determinants of Health and Hypertension Control in Adults with Medicaid. Journal of primary care & community health 2022; 13: 21501319221142426.

183. DuBay DA, Su Z, Morinelli TA, et al. Development and future deployment of a 5 years allograft survival model for kidney transplantation. Nephrology 2019; 24(8): 855–862.

184. Ghazi L, Oakes JM, MacLehose RF, Luepker RV, Osypuk TL, Drawz PE. Neighborhood Socioeconomic Status and Identification of Patients With CKD Using Electronic Health Records. American journal of kidney diseases: the official journal of the National Kidney Foundation 2021; 78(1): 57–65.e1.

185. Lange SJ, Kompaniyets L, Freedman DS, et al. Longitudinal Trends in Body Mass Index Before and During the COVID-19 Pandemic Among Persons Aged 2-19 Years – United States, 2018-2020. MMWR. Morbidity and mortality weekly report 2021; 70(37): 1278–1283.

186. Roth C, Foraker RE, Payne PRO, Embi PJ. Community-level determinants of obesity: harnessing the power of electronic health records for retrospective data analysis. BMC medical informatics and decision making 2014; 14: 36.

187. Tung EL, Wroblewski KE, Boyd K, Makelarski JA, Peek ME, Lindau ST. Police-Recorded Crime and Disparities in Obesity and Blood Pressure Status in Chicago. Journal of the American Heart Association 2018; 7(7) Published online: March 24, 2018. doi:10.1161/JAHA.117.008030.

188. Schlauch KA, Read RW, Neveux I, Lipp B, Slonim A, Grzymski JJ. The Impact of ACEs on BMI: An Investigation of the Genotype-Environment Effects of BMI. Frontiers in genetics 2022; 13: 816660.

189. Boone-Heinonen J, Tillotson CJ, O’Malley JP, et al. Characterizing a “Big Data” Cohort of Over 200,000 Low-Income U.S. Infants and Children for Obesity Research: The ADVANCE Early Life Cohort. Maternal and child health journal 2017; 21(3): 421–431.

190. Nemirovsky DR, Klose C, Wynne M, et al. Role of Race and Insurance Status in Prostate Cancer Diagnosis-to-Treatment Interval. Clinical genitourinary cancer 2023; 21(3): e198– e203.

191. Chakravarthy R, Stallings SC, Velez Edwards DR, et al. Determinants of stage at diagnosis of HPV-related cancer including area deprivation and clinical factors. Journal of public health 2022; 44(1): 18–27.

192. Cui W, Finkelstein J. Using EHR Data to Identify Social Determinants of Health Affecting Disparities in Cancer Survival. Studies in health technology and informatics 2022; 290: 967–971.

193. Byhoff E, Guardado R, Xiao N, Nokes K, Garg A, Tripodis Y. Association of Unmet Social Needs with Chronic Illness: A Cross-Sectional Study. Population health management 2022; 25(2): 157–163.

194. Kurani SS, McCoy RG, Lampman MA, et al. Association of Neighborhood Measures of Social Determinants of Health With Breast, Cervical, and Colorectal Cancer Screening Rates in the US Midwest. JAMA network open 2020; 3(3): e200618.

195. Halbert CH, Jefferson M, Allen CG, et al. Racial Differences in Patient Portal Activation and Research Enrollment Among Patients With Prostate Cancer. JCO clinical cancer informatics 2021; 5: 768–774.

196. Adie Y, Kats DJ, Tlimat A, et al. Neighborhood Disadvantage and Lung Cancer Incidence in Ever-Smokers at a Safety Net Health-Care System: A Retrospective Study. Chest 2020; 157(4): 1021–1029.

197. Edwards CV, Sheikh AR, Dennis MJ, et al. The impact of substance use on health care utilization, treatment, and outcomes in patients with non-small cell lung cancer. Journal of thoracic disease 2022; 14(10): 3865–3875.

198. Choi HY, Graetz I, Shaban-Nejad A, et al. Social Disparities of Pain and Pain Intensity Among Women Diagnosed With Early Stage Breast Cancer. Frontiers in oncology 2022; 12: 759272.

199. Sathe C, Accordino MK, DeStephano D, Shah M, Wright JD, Hershman DL. Social determinants of health and CDK4/6 inhibitor use and outcomes among patients with metastatic breast cancer. Breast cancer research and treatment 2023; 200(1): 85–92.

200. Hayes-Larson E, Ikesu R, Fong J, et al. Association of Education With Dementia Incidence Stratified by Ethnicity and Nativity in a Cohort of Older Asian American Individuals. JAMA network open 2023; 6(3): e231661.

201. Alemi F, Avramovic S, Renshaw KD, Kanchi R, Schwartz M. Relative accuracy of social and medical determinants of suicide in electronic health records. Health services research 2020; 55 Suppl 2(Suppl 2): 833–840.

202. Blosnich JR, Montgomery AE, Dichter ME, et al. Social Determinants and Military Veterans’ Suicide Ideation and Attempt: a Cross-sectional Analysis of Electronic Health Record Data. Journal of general internal medicine 2020; 35(6): 1759–1767.

203. Liu J, Hung P, Liang C, et al. Multilevel determinants of racial/ethnic disparities in severe maternal morbidity and mortality in the context of the COVID-19 pandemic in the USA: protocol for a concurrent triangulation, mixed-methods study. BMJ open 2022; 12(6): e062294.

204. Freeman C, Stanhope KK, Wichmann H, Jamieson DJ, Boulet SL. Neighborhood deprivation and severe maternal morbidity in a medicaid-Insured population in Georgia. The journal of maternal-fetal & neonatal medicine: the official journal of the European Association of Perinatal Medicine, the Federation of Asia and Oceania Perinatal Societies, the International Society of Perinatal Obstetricians 2022; 35(25): 10110–10115.

205. Cottrell EK, O’Malley JP, Dambrun K, et al. The Impact of Social and Clinical Complexity on Diabetes Control Measures. Journal of the American Board of Family Medicine: JABFM 2020; 33(4): 600–610.

206. Kind A. Neighborhood Atlas – Home. [cited January 29, 2024]. (https://www.neighborhoodatlas.medicine.wisc.edu/).

207. Ryan Powell W, Sheehy AM, Kind AJH. The Area Deprivation Index Is The Most Scientifically Validated Social Exposome Tool Available For Policies Advancing Health Equity. Health Affairs Forefront Published online: doi:10.1377/forefront.20230714.676093.

208. Social Deprivation Index (SDI). 2021 [cited January 29, 2024]. (https://www.graham-center.org/maps-data-tools/social-deprivation-index.html).

209. CDC/ATSDR Social Vulnerability Index. 2024 [cited January 29, 2024]. (https://www.atsdr.cdc.gov/placeandhealth/svi/index.html).

210. Gordon NP, Banegas MP, Tucker-Seeley RD. Racial-ethnic differences in prevalence of social determinants of health and social risks among middle-aged and older adults in a Northern California health plan. PloS one 2020; 15(11): e0240822.

211. Jordan CAL, Alizadeh F, Ramirez LS, Kimbro R, Lopez KN. Obesity in Pediatric Congenital Heart Disease: The Role of Age, Complexity, and Sociodemographics. Pediatric cardiology 2023; 44(6): 1251–1261.

212. Ahmad TR, Kong AW, Turner ML, et al. Socioeconomic Correlates of Keratoconus Severity and Progression. Cornea 2023; 42(1): 60–65.

213. Mansfield LN, Chung RJ, Silva SG, Merwin EI, Gonzalez-Guarda RM. Social determinants of human papillomavirus vaccine series completion among U.S. adolescents: A mixed-methods study. SSM – population health 2022; 18: 101082.

214. Islam JY, Madhira V, Sun J, et al. Racial disparities in COVID-19 test positivity among people living with HIV in the United States. International journal of STD & AIDS 2022; 33(5): 462–466.

215. Thompson HM, Sharma B, Smith DL, et al. Machine Learning Techniques to Explore Clinical Presentations of COVID-19 Severity and to Test the Association With Unhealthy Opioid Use: Retrospective Cross-sectional Cohort Study. JMIR public health and surveillance 2022; 8(12): e38158.

216. Chavez LJ, Tyson DP, Davenport MA, Kelleher KJ, Chisolm DJ. Social Needs as a Risk Factor for Positive Postpartum Depression Screens in Pediatric Primary Care. Academic pediatrics 2023; 23(7): 1411–1416.

217. Kapoor N, Lynch EA, Lacson R, et al. Predictors of Completion of Clinically Necessary Radiologist-Recommended Follow-Up Imaging: Assessment Using an Automated Closed-Loop Communication and Tracking Tool. AJR. American journal of roentgenology 2023; 220(3): 429–440.

218. Cobert J, Lantos PM, Janko MM, et al. Geospatial Variations and Neighborhood Deprivation in Drug-Related Admissions and Overdoses. Journal of urban health: bulletin of the New York Academy of Medicine 2020; 97(6): 814–822.

219. Egede LE, Walker RJ, Linde S, et al. Nonmedical Interventions For Type 2 Diabetes: Evidence, Actionable Strategies, And Policy Opportunities. Health affairs 2022; 41(7): 963– 970.

220. Eakin M, Singleterry V, Wang E, Brown I, Saynina O, Walker R. Effects of California’s New Patient Homelessness Screening and Discharge Care Law in an Emergency Department. Cureus 2023; 15(2): e35534.

221. Sokan O, Stryckman B, Liang Y, et al. Impact of a mobile integrated healthcare and community paramedicine program on improving medication adherence in patients with heart failure and chronic obstructive pulmonary disease after hospital discharge: A pilot study. Explor Res Clin Soc Pharm 2022; 8: 100201.

222. Tung EL, Abramsohn EM, Boyd K, et al. Impact of a Low-Intensity Resource Referral Intervention on Patients’ Knowledge, Beliefs, and Use of Community Resources: Results from the CommunityRx Trial. Journal of general internal medicine 2020; 35(3): 815–823.

223. McCrae JS, Robinson JAL, Spain AK, Byers K, Axelrod JL. The Mitigating Toxic Stress study design: approaches to developmental evaluation of pediatric health care innovations addressing social determinants of health and toxic stress. BMC health services research 2021; 21(1): 71.

224. Messmer E, Brochier A, Joseph M, Tripodis Y, Garg A. Impact of an On-Site Versus Remote Patient Navigator on Pediatricians’ Referrals and Families’ Receipt of Resources for Unmet Social Needs. Journal of primary care & community health 2020; 11: 2150132720924252.

225. Liu I, Cruz A, Gamcsik S, Harris SC. Reducing barriers to COVID-19 vaccine uptake through a culturally sensitive pharmacy-led patient medication education group in a behavioral health population. Journal of the American Pharmacists Association: JAPhA 2023; Published online: 2023. doi:10.1016/j.japh.2023.01.012.

226. Mathias P, Mahali LP, Agarwal S. Targeting Technology in Underserved Adults With Type 1 Diabetes: Effect of Diabetes Practice Transformations on Improving Equity in CGM Prescribing Behaviors. Diabetes care 2022; 45(10): 2231–2237.

227. Embick ER, Maeng DD, Juskiewicz I, et al. Demonstrated health care cost savings for women: findings from a community health worker intervention designed to address depression and unmet social needs. Archives of women’s mental health 2021; 24(1): 85– 92.

228. Sandel M, Hansen M, Kahn R, et al. Medical-legal partnerships: transforming primary care by addressing the legal needs of vulnerable populations. Health affairs 2010; 29(9): 1697– 1705.

229. Krist AH, O’Loughlin K, Woolf SH, et al. Enhanced care planning and clinical-community linkages versus usual care to address basic needs of patients with multiple chronic conditions: a clinician-level randomized controlled trial. Trials 2020; 21(1): 517.

230. Hsu C, Hertel E, Johnson E, et al. Evaluation of the Learning to Integrate Neighborhoods and Clinical Care Project: Findings from Implementing a New Lay Role into Primary Care Teams to Address Social Determinants of Health. The Permanente journal 2018; 22 Published online: 2018. doi:10.7812/tpp/18-101.

231. Berkowitz SA, Hulberg AC, Placzek H, et al. Mechanisms Associated with Clinical Improvement in Interventions That Address Health-Related Social Needs: A Mixed-Methods Analysis. Population health management 2019; 22(5): 399–405.

232. Henize AW, Beck AF, Klein MD, Adams M, Kahn RS. A Road Map to Address the Social Determinants of Health Through Community Collaboration. Pediatrics 2015; 136(4): e993– 1001.

233. Gyamfi J, Cooper C, Barber A, et al. Needs assessment and planning for a clinic-community-based implementation program for hypertension control among blacks in New York City: a protocol paper. Implement Sci Commun 2022; 3(1): 96.

234. Emmert-Aronson B, Grill KB, Trivedi Z, Markle EA, Chen S. Group Medical Visits 2.0: The Open Source Wellness Behavioral Pharmacy Model. Journal of alternative and complementary medicine 2019; 25(10): 1026–1034.

235. Taber KA, Williams JN, Huang W, et al. Use of an Integrated Care Management Program to Uncover and Address Social Determinants of Health for Individuals With Lupus. ACR Open Rheumatol 2021; 3(5): 305–311.

236. Battaglia TA, Freund KM, Haas JS, et al. Translating research into practice: Protocol for a community-engaged, stepped wedge randomized trial to reduce disparities in breast cancer treatment through a regional patient navigation collaborative. Contemporary clinical trials 2020; 93: 106007.

237. Loo S, Anderson E, Lin JG, et al. Evaluating a social risk screening and referral program in an urban safety-net hospital emergency department. J Am Coll Emerg Physicians Open 2023; 4(1): e12883.

238. Zuckerman AD, Mourani J, Smith A, et al. 2022 ASHP Survey of Health-System Specialty Pharmacy Practice: Clinical Services. American journal of health-system pharmacy: AJHP: official journal of the American Society of Health-System Pharmacists 2023; Published online: 2023. doi:10.1093/ajhp/zxad064.

239. Dagenais S, Russo L, Madsen A, Webster J, Becnel L. Use of Real-World Evidence to Drive Drug Development Strategy and Inform Clinical Trial Design. Clinical pharmacology and therapeutics 2022; 111(1): 77–89.

240. Alcaraz KI, Wiedt TL, Daniels EC, Yabroff KR, Guerra CE, Wender RC. Understanding and addressing social determinants to advance cancer health equity in the United States: A blueprint for practice, research, and policy. CA: a cancer journal for clinicians 2020; 70(1): 31–46.

241. Fox A. Scaling SDOH initiatives with analytics and coordinated workflows. Healthcare IT News. 2023 [cited January 29, 2024]. (https://www.healthcareitnews.com/news/scaling-sdoh-initiatives-analytics-and-coordinated-workflows).

242. Yan AF, Chen Z, Wang Y, et al. Effectiveness of Social Needs Screening and Interventions in Clinical Settings on Utilization, Cost, and Clinical Outcomes: A Systematic Review. Health Equity 2022; 6(1): 454–475.

243. Sawatzky R, Kwon J-Y, Barclay R, et al. Implications of response shift for micro-, meso-, and macro-level healthcare decision-making using results of patient-reported outcome measures. Quality of life research: an international journal of quality of life aspects of treatment, care and rehabilitation 2021; 30(12): 3343–3357.

244. Barasa EW, Molyneux S, English M, Cleary S. Setting healthcare priorities in hospitals: a review of empirical studies. Health policy and planning 2015; 30(3): 386–396.

245. McKneally MF, Dickens BM, Meslin EM, Singer PA. Bioethics for clinicians: 13. Resource allocation. CMAJ: Canadian Medical Association journal = journal de l’Association medicale canadienne 1997; 157(2): 163–167.

246. Midboe AM, Gray C, Cheng H, Okwara L, Gale RC. Implementation of health-focused interventions in vulnerable populations: protocol for a scoping review. BMJ open 2020; 10(7): e036937.

247. Pfaff K, Krohn H, Crawley J, et al. The little things are big: evaluation of a compassionate community approach for promoting the health of vulnerable persons. BMC public health 2021; 21(1): 2253.

248. Small N, Ong BN, Lewis A, et al. Co-designing new tools for collecting, analysing and presenting patient experience data in NHS services: working in partnership with patients and carers. Research involvement and engagement 2021; 7(1): 85.

249. Silvola S, Restelli U, Bonfanti M, Croce D. Co-Design as Enabling Factor for Patient-Centred Healthcare: A Bibliometric Literature Review. ClinicoEconomics and outcomes research: CEOR 2023; 15: 333–347.

250. Holmes JH, Beinlich J, Boland MR, et al. Why Is the Electronic Health Record So Challenging for Research and Clinical Care? Methods of information in medicine 2021; 60(1-02): 32–48.

251. Heacock ML, Lopez AR, Amolegbe SM, et al. Enhancing Data Integration, Interoperability, and Reuse to Address Complex and Emerging Environmental Health Problems. Environmental science & technology 2022; 56(12): 7544–7552.

252. Health and Human Services Department. Health Data, Technology, and Interoperability: Certification Program Updates, Algorithm Transparency, and Information Sharing. Federal Register. 2023, pp. 23746–23917.

253. Social determinants of health. [cited January 31, 2024]. (https://www.healthit.gov/health-equity/social-determinants-health).

254. Guevara M, Chen S, Thomas S, et al. Large language models to identify social determinants of health in electronic health records. NPJ digital medicine 2024; 7(1): 6.

255. Clusmann J, Kolbinger FR, Muti HS, et al. The future landscape of large language models in medicine. Communication & medicine 2023; 3(1): 141.

